# iQC: machine-learning-driven prediction of surgical procedure uncovers systematic confounds of cancer whole slide images in specific medical centers

**DOI:** 10.1101/2023.09.19.23295798

**Authors:** Andrew J. Schaumberg, Michael S. Lewis, Ramin Nazarian, Ananta Wadhwa, Nathanael Kane, Graham Turner, Purushotham Karnam, Poornima Devineni, Nicholas Wolfe, Randall Kintner, Matthew B. Rettig, Beatrice S. Knudsen, Isla P. Garraway, Saiju Pyarajan

## Abstract

**Problem:** The past decades have yielded an explosion of research using artificial intelligence for cancer detection and diagnosis in the field of computational pathology. Yet, an often unspoken assumption of this research is that a glass microscopy slide faithfully represents the underlying disease. Here we show systematic failure modes may dominate the slides digitized from a given medical center, such that neither the whole slide images nor the glass slides are suitable for rendering a diagnosis.

**Methods:** We quantitatively define high quality data as a set of whole slide images where the type of surgery the patient received may be accurately predicted by an automated system such as ours, called “iQC”. We find iQC accurately distinguished biopsies from nonbiopsies, e.g. prostatectomies or transurethral resections (TURPs, a.k.a. prostate chips), only when the data qualitatively appeared to be high quality, e.g. vibrant histopathology stains and minimal artifacts.

Crucially, prostate needle biopsies appear as thin strands of tissue, whereas prostatectomies and TURPs appear as larger rectangular blocks of tissue. Therefore, when the data are of high quality, iQC (i) accurately classifies pixels as tissue, (ii) accurately generates statistics that describe the distribution of tissue in a slide, and (iii)accurately predicts surgical procedure from said statistics.

We additionally compare our “iQC” to “HistoQC”, both in terms of how many slides are excluded and how much tissue is identified in the slides.

**Results:** While we do not control any medical center’s protocols for making or storing slides, we developed the iQC tool to hold all medical centers and datasets to the same objective standard of quality. We validate this standard across five Veterans Affairs Medical Centers (VAMCs) and the Automated Gleason Grading Challenge (AGGC) 2022 public dataset. For our surgical procedure prediction task, we report an Area Under Receiver Operating Characteristic (AUROC) of 0.9966-1.000 at the VAMCs that consistently produce high quality data and AUROC of 0.9824 for the AGGC dataset. In contrast, we report an AUROC of 0.7115 at the VAMC that consistently produced poor quality data. An attending pathologist determined poor data quality was likely driven by faded histopathology stains and protocol differences among VAMCs. Corroborating this, iQC’s novel stain strength statistic finds this institution has significantly weaker stains (*p <* 2.2 *×* 10^*−*16^, two-tailed Wilcoxon rank-sum test) than the VAMC that contributed the most slides, and this stain strength difference is a large effect (Cohen’s *d* = 1.208).

In addition to accurately detecting the distribution of tissue in slides, we find iQC recommends only 2 of 3736 VAMC slides (0.005%) be reviewed for inadequate tissue. With its default configuration file, HistoQC excluded 89.9% of VAMC slides because tissue was not detected in these slides. With our customized configuration file for HistoQC, we reduced this to 16.7% of VAMC slides. Strikingly, the default configuration of HistoQC included 94.0% of the 1172 prostate cancer slides from The Cancer Genome Atlas (TCGA), which may suggest HistoQC defaults were calibrated against TCGA data but this calibration did not generalize well to non-TCGA datasets. For VAMC and TCGA, we find a negligible to small degree of agreement in the include/exclude status of slides, which may suggest iQC and HistoQC are not equivalent.

**Conclusion:** Our surgical procedure prediction AUROC may be a quantitative indicator positively associated with high data quality at a medical center or for a specific dataset. We find iQC accurately identifies tissue in slides and excludes few slides, unless the data are poor quality. To produce high quality data, we recommend producing slides using robotics or other forms of automation whenever possible. We recommend scanning slides digitally before the glass slide has time to develop signs of age, e.g faded stains and acrylamide bubbles. We recommend using high-quality reagents to stain and mount slides, which may slow aging. We recommend protecting stored slides from ultraviolet light, from humidity, and from changes in temperature. To our knowledge, iQC is the first automated system in computational pathology that validates data quality against objective evidence, e.g. surgical procedure data available in the EHR or LIMS, which requires zero efforts or annotations from anatomic pathologists. Please see https://github.com/schaumba/iqc and https://doi.org/10.17605/OSF.IO/AVD3Z for instructions and updates.

## 1 Introduction

Over the past several years in the field of computational pathology[1], automated methods to assess data quality have tended to focus on excluding confounded regions or measuring the negative impact of poor quality data. The field has benefited from specialized tools such as tissue fold detection[2], blur detection[3], pen detection[4], and fragment detection[5] – as well as general frameworks for quality control[6, 7]. The negative effect of whole slide image artifacts on downstream computational pathology methods has been benchmarked[8, 9], with some reports of specific normalizations improving task performance under specific artifacts[10, 11].

If an automated system is not used to segment out artifacts or other confounds that would lower data quality, computational pathology pipelines may implicitly or explicitly have methods to exclude artifacts. For instance, our earlier work to predict SPOP mutation in prostate cancer was engineered to focus on diagnostically salient regions enriched in predicted subtypes of nuclei, which implicitly avoids pen and background[12]. Later, in what would form the basis of Paige Prostate, Campanella and colleagues used weakly supervised learning and a recurrent neural network to identify suspect foci of cancer in whole slide images, which explicitly used Otsu’s method[13] to exclude background and through machine learning at scale may implicitly learn to avoid some artifacts[14]. Shortly thereafter, Lu and colleagues used weakly supervised learning with an attention mechanism for renal cancer subtyping, which explicitly used thresholding to exclude background, while their attention mechanism was shown to exclude normal morphology and some artifacts[15].

Unfortunately, in a large independent validation, Perincheri and colleagues noted “Areas for improvement were identified in Paige Prostate’s handling of poor quality scans”, which may suggest weakly supervised learning may benefit from rigorous quality control as a preprocessing step[16]. Quality control may flag poor quality slides for manual review, exclude artifacts, or take other actions before Paige Prostate or other downstream processing occurs.

Inspired by our early work that noted different surgical procedures (Fig 1A2,A3) may impact the distribution of tissue in a slide and deep learning performance[17], we developed the hypothesis underlying our iQC tool. Specifically, for high quality data, a quality control system should be able to accurately count the number of tissue pixels in a slide, describe the distribution of tissue in a slide, and therefore predict what kind of surgical procedure was used to excise the tissue present in the slide (Fig 2A1,C1,D1,F1,Q1 are biopsy examples versus Fig 2B1,L2,N1,O1,P1 are nonbiopsy examples). Thus for poor quality data, our hypothesis is that tissue may not be accurately measured and surgical procedure may not be accurately predicted.

**Fig 1.**
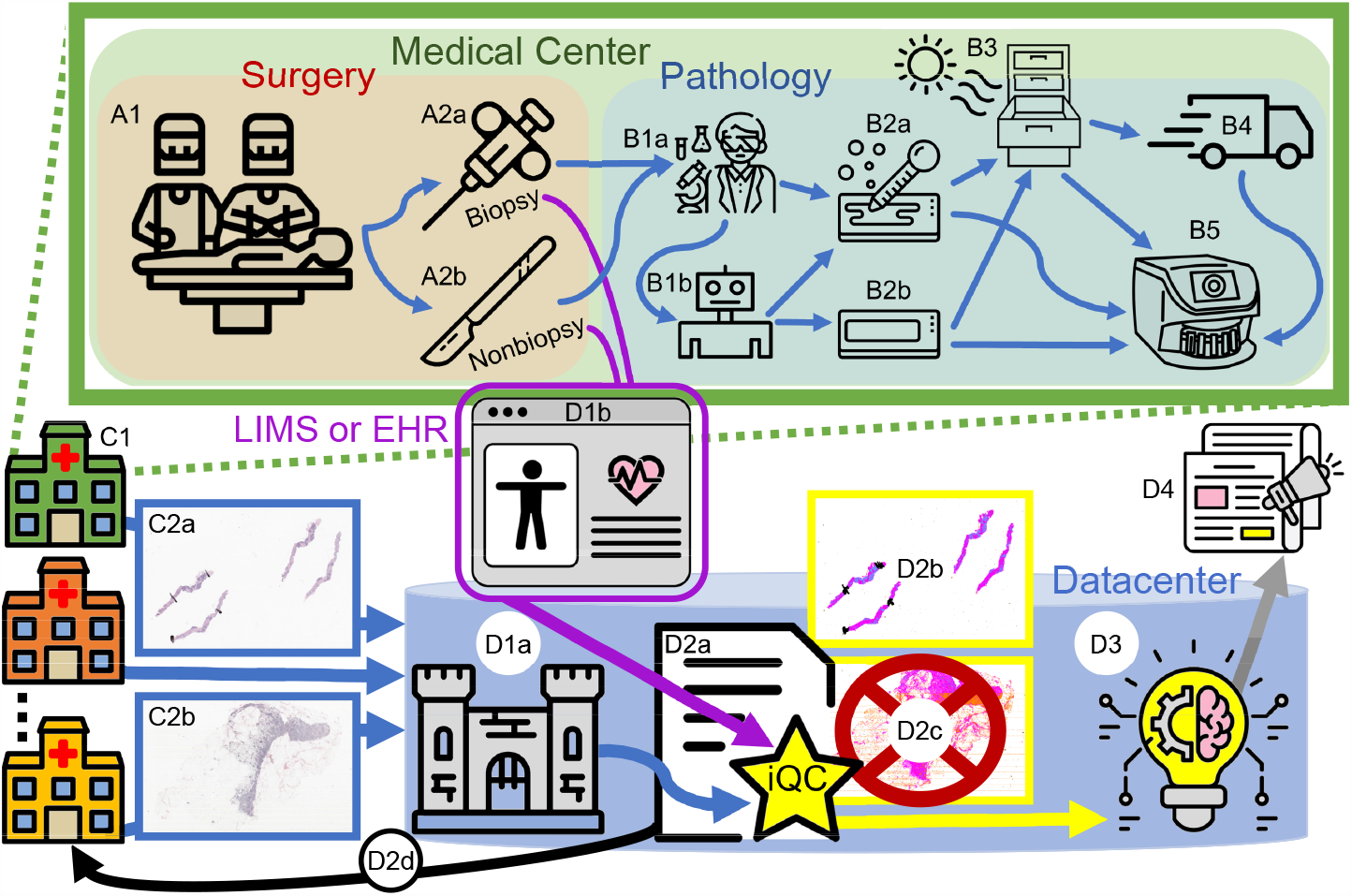
Our study’s workflow (Sec S1.9), from a patient’s surgery (A1), through iQC’s quality control (D2a-c), to downstream computational research (D3) and results (D4). **A1**: Tissue is surgically excised from the patient. **A2a**: Tissue may be excised as a needle biopsy, which removes a thin ribbon of tissue (Fig 2A1). **A2b**: Alternatively, tissue may be excised as a “nonbiopsy”, e.g. a prostatectomy (Fig 2E1) or a transurethral resection of prostate (TURP). **B1a**: A pathologists’ assistant receives the tissue. **B1b**: The pathologists’ assistant may use a machine to prepare the slide. **B2a**: A slide may be prepared by hand or by machine, using stains and *mounting solution*. **B2b**: Alternatively, a machine may prepare the slide using stains and a lower-cost *mounting tape*. **B3**: A prepared slide is stored, e.g. in a cabinet in the pathology department or medical center. Slides age here and may be subject to changes in heat, humidity, ultraviolet light, etc. Aging rates may depend on slide preparation. **B4**: If the pathology department does not have a whole slide scanner, the department ships the slides to a different medical center that has one. **B5**: A whole slide scanner takes a high-resolution picture of the entire slide. This picture is a whole slide image. **C1**: Several medical centers in parallel generate whole slide images. **C2a**: Each medical center sends a mix of whole slide images, which may include biopsies (c.f. A2a). **C2b**: Nonbiopsy images may be sent in the mix (c.f. A2b). **D1a**: Images are collected and organized in the datacenter. **D1b**: The Laboratory Information Management System (LIMS) tracks which slides are biopsies (e.g. C2a) and which slides are nonbiopsies (e.g. C2b). These biopsy/nonbiopsy metadata are used to train iQC (D2a) to predict from the image the corresponding surgical procedure (biopsy vs nonbiopsy). Alternatively, surgical procedure may be tracked in the Electronic Health Record (EHR). **D2a**: iQC subjects all whole slide images to quality control and generates many statistics to describe each image. **D2b**: iQC generates a mask image to describe each pixel (c.f. C2a), and in this case passes biopsy samples on to downstream studies (D3). **D2c**: iQC may exclude poor quality slides or exclude nonbiopsies (c.f. C2b) from downstream studies (D3), if the studies are intended to be of biopsies only. **D2d**: iQC generates reports for medical centers to act upon, e.g. remaking poor quality slides. **D3**: Downstream computational studies may occur on high-quality whole slide images of the appropriate surgical procedure type, i.e. biopsies. **D4**: Studies may lead to publication.

**Fig 2.**
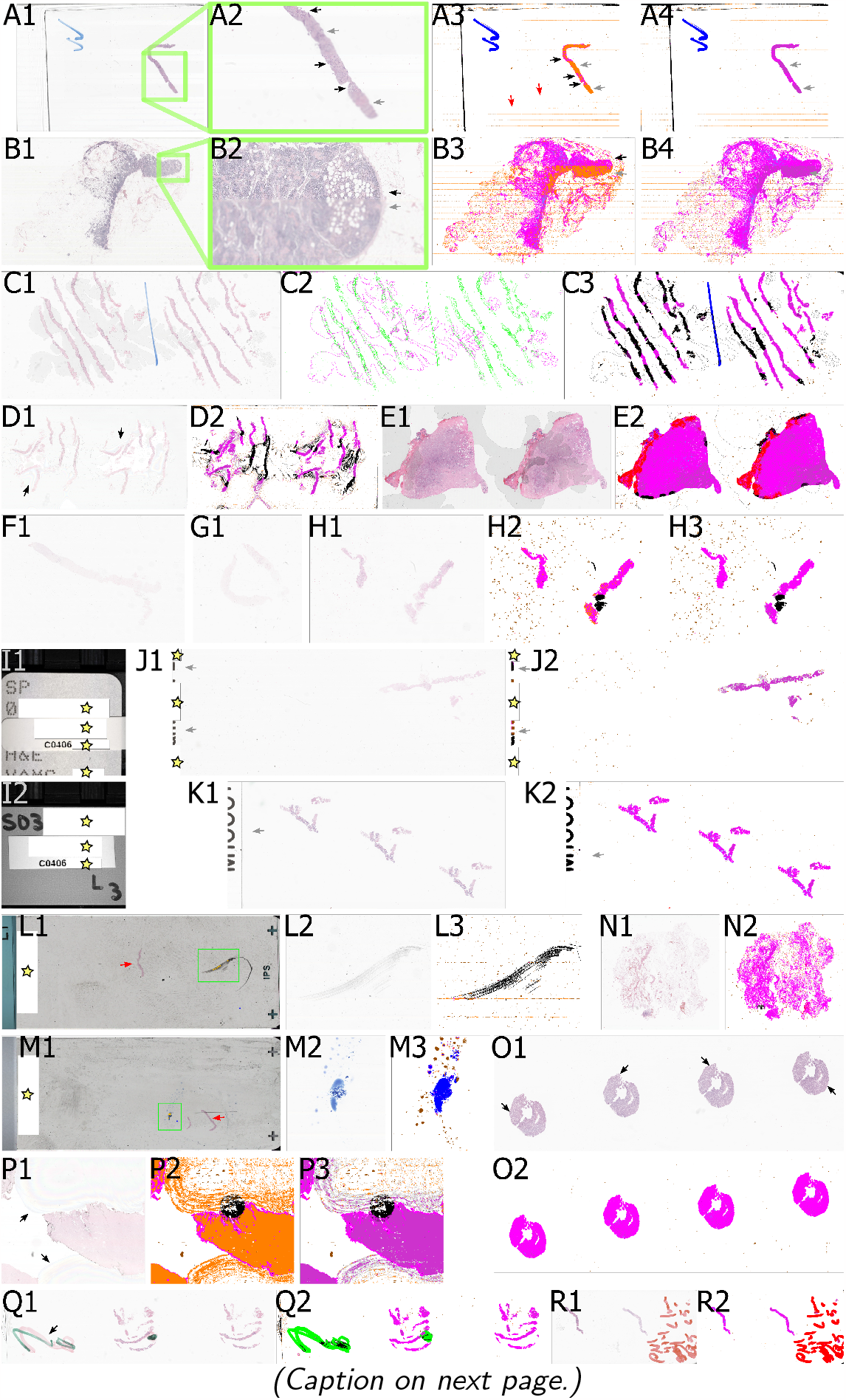
Representative histopathology images in our study. **A1**: a whole slide image of a prostate needle biopsy at low magnification. **A2**: High magnification of prostate needle biopsy from A1, showing horizontal bands of systematic blur (gray arrows), in contrast to bands of visually sharp pixels where glands are visible (black arrows). **A3**: iQC’s quality control mask 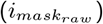 showing the type of each pixel – background pixels are in white, blue pen pixels (which draw two blue check marks rotated 90°) are in blue, the edge of the slide is in black, tissue pixels are in magenta (black arrows), “suspect” tissue pixels are orange (gray arrows), and “suspect” pixels that form horizontal bands (red arrows) may suggest the quality of this whole slide image suffers from systematic blur. **A4**: iQC’s quality control mask 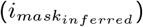 shows machine learning infers “suspect” tissue pixels as tissue (dark magenta at gray arrows), so all tissue in the slide may be accurately measured for biopsy/nonbiopsy prediction. **B1**: A pelvic lymph node at low magnification where systematic blur may be difficult to perceive. **B2**: Higher magnification plainly shows a horizontal band of systematic blur (gray arrow) compared to visually sharp pixels (black arrow). **B3**: This 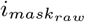 shows sharp pixels are assigned the “tissue” type as indicated in magenta (black arrow), while systematically blurred pixels have the “suspect” type as indicated in orange (gray arrow). **B4**: Machine learning infers “suspect” tissue pixels as tissue, which is shown as a dark magenta (gray arrow). **C1**: A prostate needle biopsy, where the slide shows signs of age. **C2**: iQC’s quality control mask 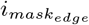 outlines in magenta these signs of age, i.e. large acrylamide bubbles from degraded mounting compound incompletely holding the coverslip to the glass slide (Sec S1.6). Tissue and pen are outlined in green. 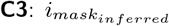 shows some tissue pixels in magenta, while other tissue pixels are shown in black, which may loosely correspond to which tissue is most confounded by bubbles. Bubble edges are shown in black. **D1**: This slide shows signs of age through refractive dispersion that causes a rainbow effect (black arrows), in addition to bubbles. **D2**: Like C3, 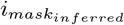 shows tissue in magenta and the most age-confounded pixels in black. **E1**: The Cancer Genome Atlas (TCGA) is a public dataset. There are bubbles throughout slide [TCGA-QU-A6IM-01Z](https://portal.gdc.cancer.gov/files/5cd4c553-e450-42e0-8129-7b1abf9cc09f), which may emphasize the value of automated quality control for public datasets. 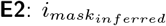 shows bubble edges or bubble-confounded regions in black, regions with blood/erythrocytes in red, and regions with blue marker in blue. **F1**,**G1**,**H1**: all these slides have faded histopathology stains. A pathologist deemed these slides unsuitable for diagnosis, corroborating iQC’s stain strength statistics (Sec S1.2.7.1) that are weak for these slides. **H2**,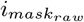 and 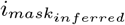, respectively, which show points of debris as brown spots, threads of debris in black, and thread-confounded tissue in black. **I1**: A whole slide image thumbnail showing identifiers such as surgical pathology number “SP…” that may be printed on the glass slide, along with other identifiers. A coded external ID “C…” may be applied as a sticker on top to redact some or all of these identifiers. We indicate our redactions to this image with stars. **I2**: A whole slide image thumbnail that shows an accession number “S03…” and a coded external ID “C…”. We remove all thumbnails from slides because no identifiers are allowed in research data. Our redactions are indicated with stars. **J1**,**J2**,**K1**,**K2**: Depending on how the glass slide is physically aligned during scanning, text or potentially identifiers on the slide (see I1,I2) may be scanned in the whole slide image at high resolution (at gray arrows, stars for redactions). iQC flags for manual review slides having such markings because patient names or other identifiers are not allowed in research data. **L1**: The thumbnail indicates a black scuff artifact was scanned at high resolution (green box) – missing the prostate needle biopsy (red arrow). **L2**,**L3**: There is no human tissue scanned at high resolution in this slide, only the black artifact. **M1**: The thumbnail indicates a blue pen mark was scanned at high resolution (green box) – missing the prostate needle biopsy (red arrow). **M2**,**M3**: There is no human tissue scanned at high resolution in this slide, only the blue artifact. **N1**,**N2**: Iliac bone in our dataset, due to metastasis. **O1**,**O2**: Colon polypectomy in our dataset, with colonic crypts visible (black arrows). **P1**: Slide with faded stain and more extensive refractive dispersion (black arrows) than D1. **P2**: Due to faded stain and slide age, many pixels are have the “suspect” type in 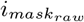 (orange). **P3**: Underlining the importance of iQC’s machine learning to infer pixel types, these pixels are re-typed as tissue (dark magenta) in 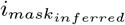. Other suspect pixels are inferred as background (gray). **Q1**,**Q2**: Green pen over red pen (black arrow) typed as green or black in 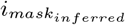. **R1**,**R2**: iQC detects red pen.

## 2 Results

### 2.1 Batch effect of poor quality data detected

We found a batch effect, where a subset of data accounted for most slides in iQC’s “fail_all_tissue” category (Fig 3A1). iQC defines ten quality control categories (Sec S1.1). Specifically, we found Institution *β* accounted for most of the “fail_all_tissue” slides (Fig 3A2). An anatomic pathologist recommended all slides from Institution *β* fail quality control and be remade (Figs 3B2 and 1D2d).

**Fig 3.**
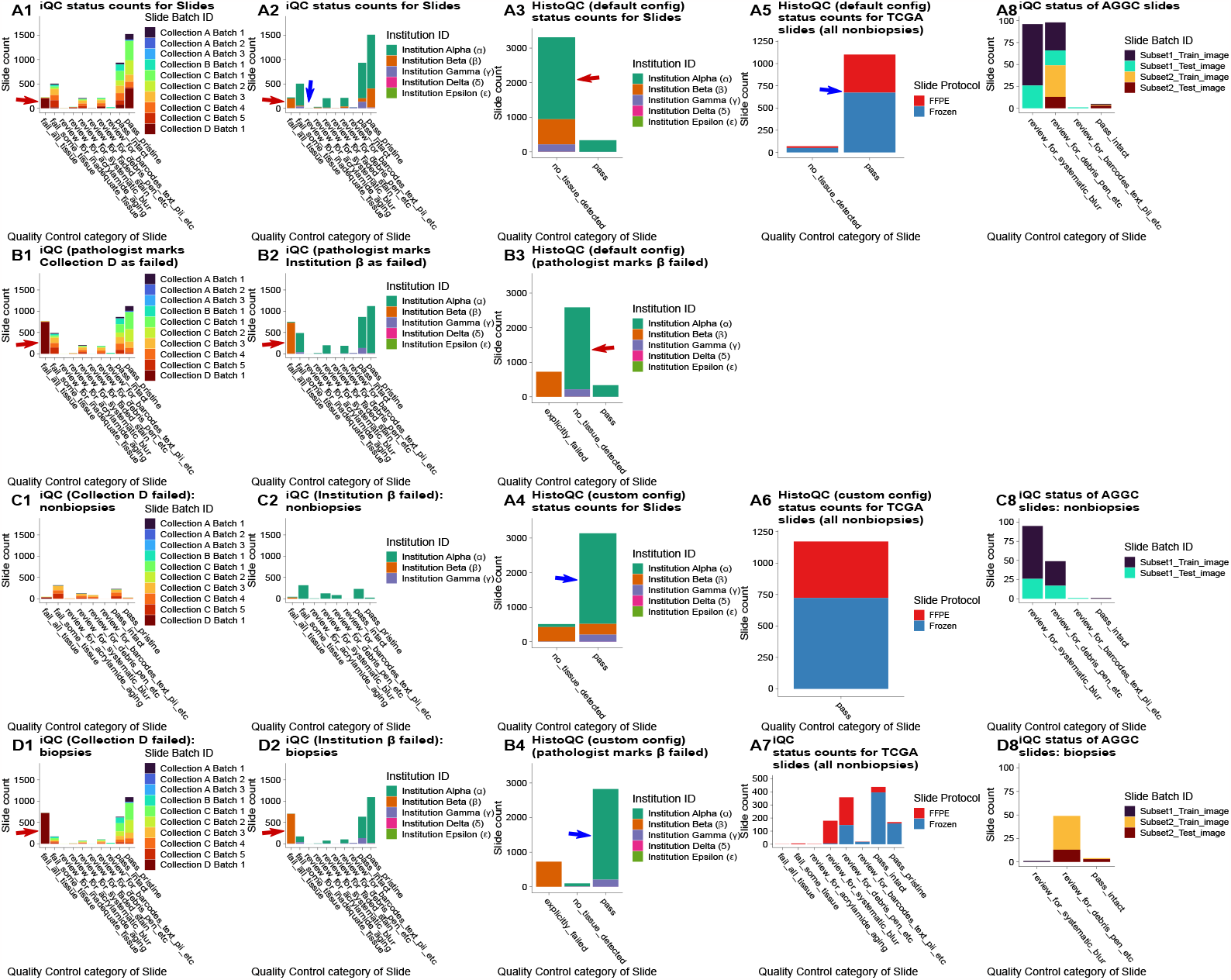
iQC and HistoQC quality control category counts for slides in VAMC, TCGA, and AGGC datasets. **A1**: Initial data collection. A “collection” is a set of data, e.g. all slides from a conference room or a prior study. A “batch” corresponds to a subset of slides, e.g. the first box of slides from the conference room. The next box is batch 2. We noticed “Collection D Batch 1” accounted for most slides in iQC’s “fail_all_tissue” category (red arrow), which indicates these slides have faded stain, so the entire slide is unsuitable for diagnosis. **A2**: Coloring A1 according to the VAMC/Institution that made the slide, Institution Beta (*β*) accounts for most “fail_all_tissue” slides, which is a batch effect (red arrow). iQC excludes 227 slides as “fail_all_tissue”, 505 slides as “fail some tissue”. **A3**: With the default configuration file, HistoQC excludes (89.9%) of slides as “no tissue detected” (red arrow). For comparison, iQC only flags 2 slides as “review for inadequate tissue” (panel A2’s blue arrow). **A4**: With our customized configuration file, HistoQC excludes (16.7%) of slides as “no tissue detected” (blue arrow), which is much better than the default configuration’s 89.9% (panel A3’s red arrow). For comparison, iQC only excludes 227 slides (6.2%) as “fail_all_tissue” (panel A2’s red arrow). **A5**: All but 70 of 1172 (94.0%) TCGA slides pass (blue arrow) HistoQC_default configuration_, which may suggest HistoQC’s defaults are calibrated against TCGA. All TCGA slides are nonbiopsies. **A6**: HistoQC_custom configuration_ includes all TCGA slides. **A7**: iQC includes all but 7 of 1172 (99.4%) of TCGA slides. iQC recommended “review for inadequate tissue” for zero slides, while HistoQC excluded 70 slides as “no tissue detected” (c.f. A5). **A8**: iQC excludes zero AGGC slides, suggesting this dataset is high quality[18]. Most slides are “review…”, which is a warning. Many slides have tissue debris. **B1**,**B2**: A pathologist reviewed slides from Institution *β*. He concluded they were poor quality and unsuitable for diagnosis. He recommended all slides from Institution *β* be excluded (red arrow). **B3**,**B4**: HistoQC’s corresponding exclusion status for Institution *β* slides is “explicitly failed”. HistoQC_default_ does not detect tissue in most slides (B3 red arrow), but HistoQC_custom_ recovers most of these slides (B4 blue arrow). **C1**,**C2**,**C8**: Quality control categories for slides iQC predicted to be nonbiopsies, e.g. prostatectomies or TURPs. **D1**,**D2**,**D8**: Quality control categories for slides iQC predicted to be biopsies of any sort, e.g. prostate needle biopsies or liver biopsies. For us, HistoQC did not accept the AGGC TIFF file format of whole slide images (Sec S1.5).

#### 2.1.1 HistoQC recapitulates iQC batch effect identification

With HistoQC, we could recapitulate this result (Sec 2.1) and find the batch effect. While the default configuration file for HistoQC (Sec S1.5) excluded most slides as “no_tissue_detected” (Fig 3A3), we could customize the HistoQC configuration file. With our custom configuration file, HistoQC identified most “no_tissue_detected” slides were from Institution *β* (Fig 3A4).

While iQC and HistoQC agree on the batch effect, their methods to exclude slides differ (Figs 4 and 5). iQC directly detects faded histopathology stains (Sec S1.2.7.1), and marks these slides as “fail_all_tissue”. In contrast, HistoQC may not detect tissue having faded stains, in which case these slides are marked as “no_tissue_detected”.

**Fig 4.**
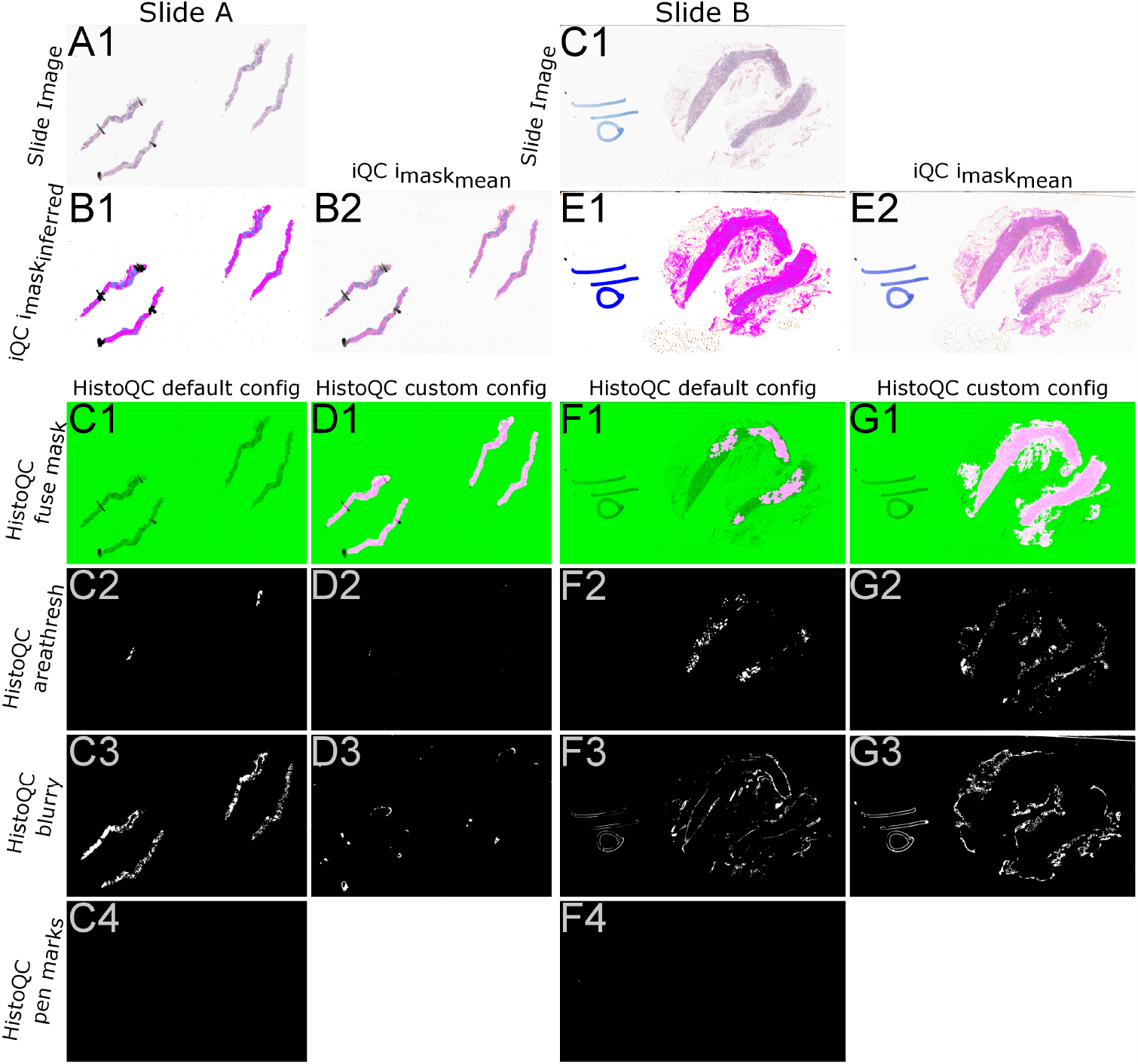
Qualitative comparison of iQC to HistoQC on VAMC slides. **A1**: A prostate needle biopsy. **B1**: iQC’s 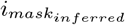 segments slide background (white), tissue (magenta), tissue with rich hematoxylin staining (cyan), black pen marks (black), and debris (brown). **B2**: iQC’s 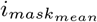 is an interpretable weighted average of the slide image (A1) and 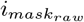 (c.f. 2A3,B3 for examples of 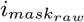), to show the mapping of slide pixels to iQC pixel types. **C1**: HistoQC excludes the entire side, per the “fuse” mask (green). **C2**: HistoQC excludes some areas for small area. **C3**: HistoQC excludes most areas as blurry. **C4**: HistoQC does not exclude any areas as pen marks. **D1**: With our custom configuration file, HistoQC segments out the tissue in the slide (pink, c.f. panel C1). We disabled pen detection in this configuration (Sec S1.5). **D2**: As expected, no small areas were excluded. **D3**: Few regions were discarded as blurry, which is better than the default configuration (panel C3). **C1**: Adipose tissue indicates this is not a biopsy (Eqn 1). **E1**: iQC’s 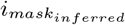segments background, tissue, blue pen, and debris. **E2**: iQC’s 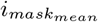 to show the mapping of pixel values to types. **F1**: HistoQC includes some tissue (magenta) and excludes the rest (green). **F2**: HistoQC excludes some tissue as small areas. **F3**: HistoQC excludes some tissue and pen as blurry. **F4**: HistoQC does not exclude any areas as pen marks. **G1**: With our custom configuration file, HistoQC segments out most of the tissue (pink) but excludes some adipose tissue that iQC detects (c.f. E1 in magenta). **G2**: HistoQC still excludes some small areas unfortunately. **G3**: HistoQC excludes more regions as blurry – perhaps worse than the default configuration (c.f. F3).

**Fig 5.**
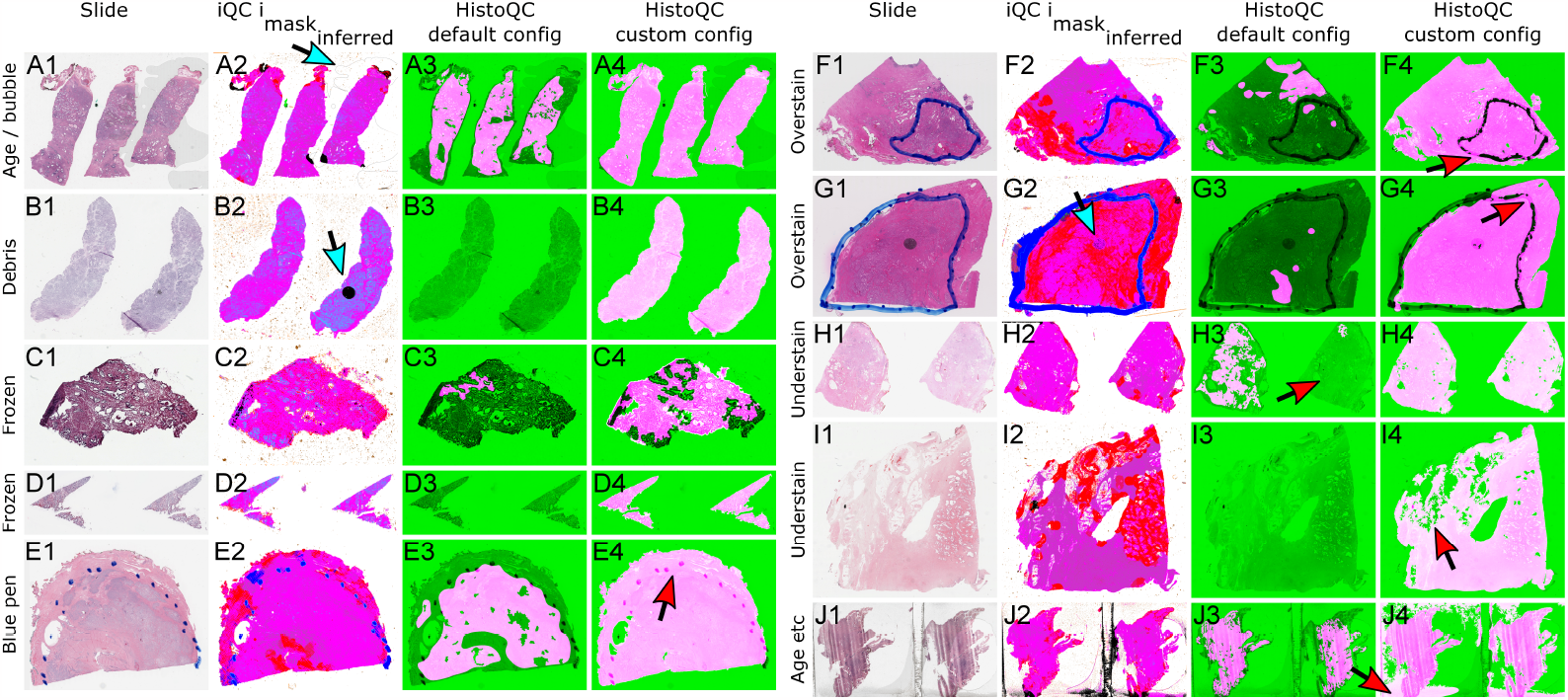
Qualitative comparison of iQC to HistoQC on TCGA slides. **A1**: An [FFPE slide (TCGA-QU-A61N)](https://portal.gdc.cancer.gov/image-viewer?filters=%7B%22content%22%3A%5B%7B%22content%22%3A%7B%22field%22%3A%22cases.case_id%22%2C%22value%22%3A%5B%22d07d20f4-dd65-4ffc-add4-01f350ca7b65%22%5D%7D%2C%22op%22%3A%22in%22%7D%5D%2C%22op%22%3A%22and%22%7D) with bubble artifacts. **A2**: iQC identifies the bubbles (cyan arrow). **A3**: HistoQC_default configuration_ excludes some tissue. **A4**: HistoQC_custom configuration_ does not exclude tissue. **B1**: An FFPE slide ([TCGA-CH-5748)](https://portal.gdc.cancer.gov/image-viewer?filters=%7B%22content%22%3A%5B%7B%22content%22%3A%7B%22field%22%3A%22cases.case_id%22%2C%22value%22%3A%5B%220af98331-9305-41e9-af55-30390016d65e%22%5D%7D%2C%22op%22%3A%22in%22%7D%5D%2C%22op%22%3A%22and%22%7D&selectedId=8b309bad-b202-450e-8f02-7d4f4106a04e) with some debris. **B2**: iQC excludes the debris as black pixels (cyan arrow). **B3**: HistoQC_default_ excludes all the tissue. **B4**: HistoQC_custom_ excludes neither tissue nor debris. **C1**: A frozen section slide ([TCGA-G9-6353)](https://portal.gdc.cancer.gov/image-viewer?filters=%7B%22content%22%3A%5B%7B%22content%22%3A%7B%22field%22%3A%22cases.case_id%22%2C%22value%22%3A%5B%22bcc94953-ce79-428e-99c1-632e42c1b9bc%22%5D%7D%2C%22op%22%3A%22in%22%7D%5D%2C%22op%22%3A%22and%22%7D&selectedId=acb2d138-90ad-4c8a-adf4-9e699dcb466e), with freezing artifacts shown as small white rips in tissue. **C2**: iQC excludes some of the tissue fold as black pixels, and includes all tissue. **C3**: HistoQC_default_ excludes most tissue. **C4**: HistoQC_custom_ includes most tissue. **D1**: A [frozen section slide (TCGA-EJ-AB27)](https://portal.gdc.cancer.gov/image-viewer?filters=%7B%22content%22%3A%5B%7B%22content%22%3A%7B%22field%22%3A%22cases.case_id%22%2C%22value%22%3A%5B%22b7b61392-c085-4e28-892f-e9b6fe6ece41%22%5D%7D%2C%22op%22%3A%22in%22%7D%5D%2C%22op%22%3A%22and%22%7D), with freezing artifacts. **D2**: iQC retains all tissue. **D3**: HistoQC_default_ excludes all tissue. **D4**: HistoQC_custom_ includes all tissue. **E1**: An [FFPE slide (TCGA-V1-A9ZR)](https://portal.gdc.cancer.gov/image-viewer?filters=%7B%22content%22%3A%5B%7B%22content%22%3A%7B%22field%22%3A%22cases.case_id%22%2C%22value%22%3A%5B%2223ebc73d-be3b-42f8-9390-bf18a21afc5c%22%5D%7D%2C%22op%22%3A%22in%22%7D%5D%2C%22op%22%3A%22and%22%7D&selectedId=28adf83e-1fb4-4dad-b0d2-95940886ebee) with blue pen dots. **E2**: iQC identifies the blue dots to exclude. **E3**: HistoQC_default_ excludes most of the blue pen and some tissue. **E4**: HistoQC_custom_ includes the blue dots (red arrow) and all tissue. **F1**: An overstained [FFPE slide (TCGA-YL-A8SL)](https://portal.gdc.cancer.gov/image-viewer?filters=%7B%22content%22%3A%5B%7B%22content%22%3A%7B%22field%22%3A%22cases.case_id%22%2C%22value%22%3A%5B%2223ebc73d-be3b-42f8-9390-bf18a21afc5c%22%5D%7D%2C%22op%22%3A%22in%22%7D%5D%2C%22op%22%3A%22and%22%7D&selectedId=28adf83e-1fb4-4dad-b0d2-95940886ebee) with blue pen marks. **F2**: iQC flags some tissue as red/eosinophilic/blood, includes the remaining tissue, and excludes blue pen. **F3**: HistoQC_default_ excludes most of the blue pen and some tissue. **F4**: HistoQC_custom_ includes all tissue and includes some blue pen marks (red arrow). **G1**: An overstained [FFPE slide (TCGA-YL-A8SH)](https://portal.gdc.cancer.gov/image-viewer?filters=%7B%22content%22%3A%5B%7B%22content%22%3A%7B%22field%22%3A%22cases.case_id%22%2C%22value%22%3A%5B%22bfb973eb-d42d-4900-a606-a381d2317555%22%5D%7D%2C%22op%22%3A%22in%22%7D%5D%2C%22op%22%3A%22and%22%7D&selectedId=f50400e0-7c1a-4e89-a9a4-966a8dc46447) with blue pen marks and a dark smudge. **G2**: iQC excludes the blue pen, excludes some of the smudge as black pixel types (cyan arrow), flags much of the tissue as red/eosinophilic/blood, and includes the rest of the tissue. **G3**: HistoQC_default_ excludes most of the tissue and slide. **G4**: HistoQC_custom_ includes all tissue and includes some blue pen marks (red arrow). **H1**: An understained [FFPE slide (TCGA-G9-6361)](https://portal.gdc.cancer.gov/image-viewer?filters=%7B%22content%22%3A%5B%7B%22content%22%3A%7B%22field%22%3A%22cases.case_id%22%2C%22value%22%3A%5B%2271d14304-ccb0-4a4c-b935-8c014ca75fd0%22%5D%7D%2C%22op%22%3A%22in%22%7D%5D%2C%22op%22%3A%22and%22%7D&selectedId=dd5c10f4-9973-43c4-83b9-20977265abff), with the section at right being slightly less stained than the section at left. **H2**: iQC includes left and right sections. **H3**: HistoQC_default_ excludes most of the less-stained section at right (red arrow), but includes most of the more-stained section at left, which may suggest HistoQC’s tissue identification partly depends on tissue staining intensity. **H4**: HistoQC_custom_ includes all tissue. **I1**: An understained [FFPE slide (TCGA-Y6-A8TL)](https://portal.gdc.cancer.gov/image-viewer?filters=%7B%22content%22%3A%5B%7B%22content%22%3A%7B%22field%22%3A%22cases.case_id%22%2C%22value%22%3A%5B%22a7045324-cb44-4b37-bbaf-641642c1d903%22%5D%7D%2C%22op%22%3A%22in%22%7D%5D%2C%22op%22%3A%22and%22%7D&selectedId=125c976b-5073-4374-9cdc-8bac77ac8ead). **I2**: iQC flags much of the tissue as red/eosinophilic/blood, and includes the rest of the tissue. **I3**: HistoQC_default_ excludes all tissue. **I4**: HistoQC_custom_ excludes some of the tissue (red arrow), but includes most. **J1**: An [FFPE slide (TCGA-TK-A8OK)](https://portal.gdc.cancer.gov/image-viewer?filters=%7B%22content%22%3A%5B%7B%22content%22%3A%7B%22field%22%3A%22cases.case_id%22%2C%22value%22%3A%5B%224fdc9f5a-7ba1-4f80-b3db-2b7cbe505217%22%5D%7D%2C%22op%22%3A%22in%22%7D%5D%2C%22op%22%3A%22and%22%7D&selectedId=38d8f020-6021-476d-9e11-ca1fdd13b949) with bubbles and other signs of age or damage. **J2**: iQC excludes most of the artifacts in black and includes most of the tissue. **J3**: HistoQC_default_ includes most of the tissue. **J4**: HistoQC_custom_ includes many artifacts (red arrow) and all of the tissue.

#### 2.1.2 iQC surgical prediction performance corresponds to data quality

After manually reviewing dozens of cases in detail from Institution *α*, we determined the equations and parameters for iQC’s surgical procedure (i.e. biopsy/nonbiopsy) predictor (Sec S1.4). In this way, iQC achieved AUROC of 0.9966 for the biopsy/nonbiopsy prediction task (Fig 6A). Testing this on all other VAMC data, we found AUROC substantially dropped to AUROC of 0.8346 (Fig 6B). Testing only on Institution *β* data, we found even lower AUROC of 0.7115 (Fig 6C). Testing on the Institutions that were neither *α* nor *β*, we found AUROC of 1.000 (Fig 6D). We concluded Institution *β* drove the drop in AUROC on non-Institution-*α* data (Fig 6B).

**Fig 6.**
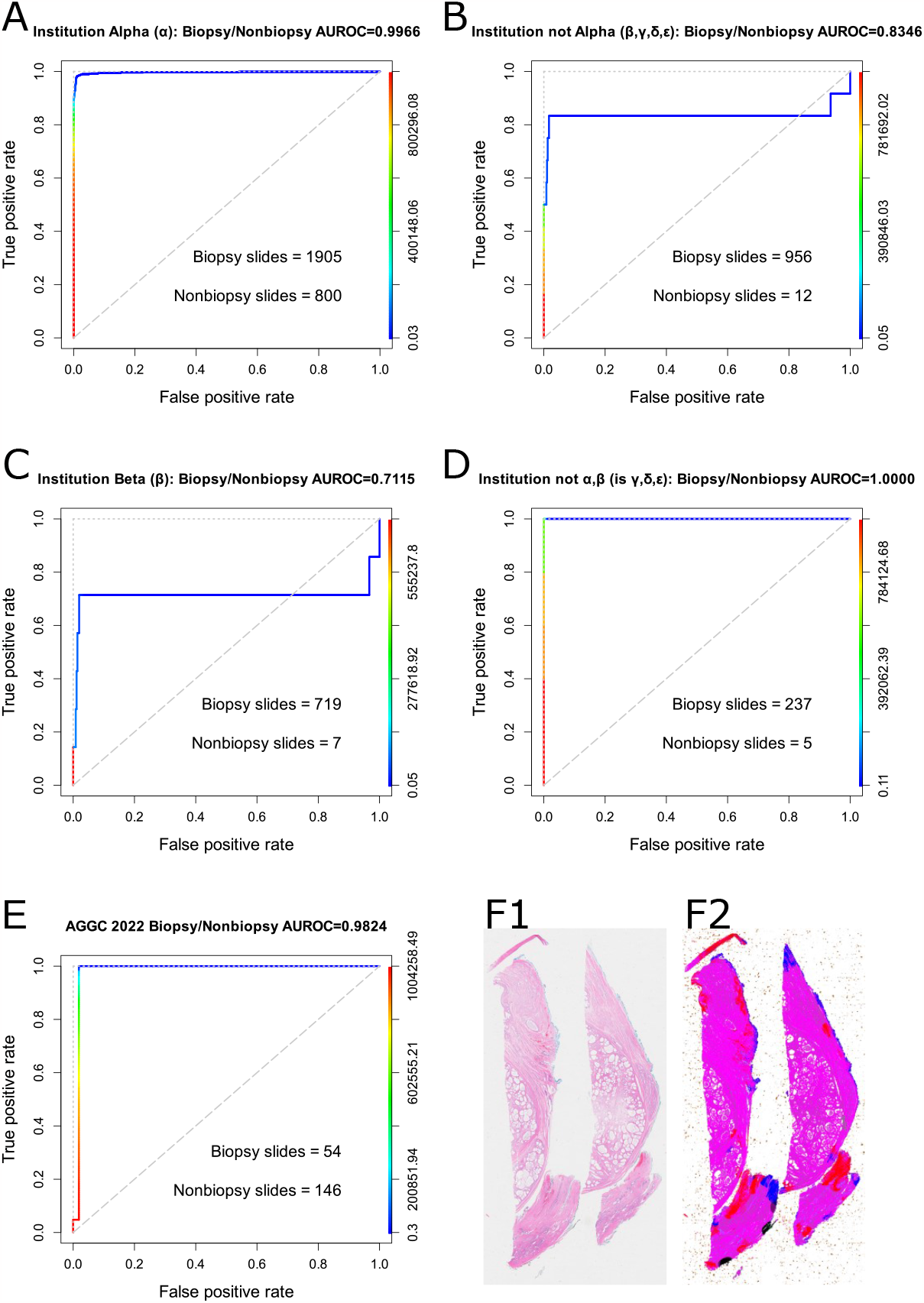
iQC biopsy/nonbiopsy prediction AUROC for VAMC and AGGC data. **A**: AUROC for Institution Alpha (*α*), which made most VAMC slides. We trained/tuned biopsy/nonbiopsy predictor on a subset of this dataset, so AUROC is high. **B**: AUROC for all institutions that are not *α*, i.e. *β, γ, δ, ϵ*. AUROC is much lower. There are few nonbiopsy slides. **C**: AUROC for *β*, which is strikingly low. We believe this is because Institution *β* provided poor quality slides that were old and had faded stain. iQC is not able to accurately identify which pixels are tissue and cannot distinguish biopsies from nonbiopsies. **D**: AUROC for other VAMCs (i.e. *γ, δ, ϵ*) is high but this is an underpowered test because there are only 5 nonbiopsies. **E**: AUROC on the public AGGC dataset[18] is high, indicating iQC’s biopsy/nonbiopsy predictor may generalize well to unseen data. **F1**,**F2**: iQC mistakenly classified this prostatectomy sample as a needle biopsy, perhaps because the distribution of tissue is long and thin, very loosely like a needle biopsy.

### 2.2 iQC standard of data quality across datasets

We next asked if iQC would serve as a standard of quality both on VAMC data and unseen high quality data from outside Veterans Affairs. In the public dataset for the Automated Gleason Grading Challenge 2022[18], we found zero slides failed iQC’s quality control, using the same quality control code, configuration, and thresholds for iQC at VAMCs (Fig 3A5). This may suggest iQC may serve as a standard of data quality across datasets without reconfiguration, but we sought corroborating evidence, below.

#### 2.2.1 iQC surgical prediction performance strong on external high quality datasets

To test how well iQC generalized to unseen data, and more specifically to non-Veteran data, we evaluated iQC’s biopsy/nonbiopsy predictor on AGGC data[18]. We found an AUROC of 0.9824 (Fig 6E). This may suggest iQC generalizes well to unseen data, external datasets, and non-Veteran data. An example of a nonbiopsy that iQC mistakes for a biopsy is shown in Figure 6F1, where the ectomy tissue is cut into a long a thin strip. Biopsies tend to be long and thin.

### 2.3 iQC’s stain strength statistic quantifies differences among VAMCs and datasets

To test if iQC could quantify differences in stain strength among VAMCs, among surgical procedures, and between VAMC and non-VAMC datasets, we considered iQC’s novel stain strength statistic for each whole slide image (Sec S1.2.7.1).

#### 2.3.1 Significant and large stain strength difference among VAMC biopsies

We found VAMC Institution *α* contributed biopsy slides that were stained significantly stronger than VAMC Institution *β*’s biopsy slides (*p* = 1.17*×*10^*−*131^, two-tailed Wilcoxon rank-sum test), and this staining effect was large (Cohen’s *d* = 1.208) (Fig 7). Corroborating our pathologist’s expert opinion that *β* slides were faded such that *β* slides were unsuitable for diagnosis (Fig 2F1,G1,H1), we suspect this large effect size as measured by Cohen’s *d* is clinically meaningful.

**Fig 7.**
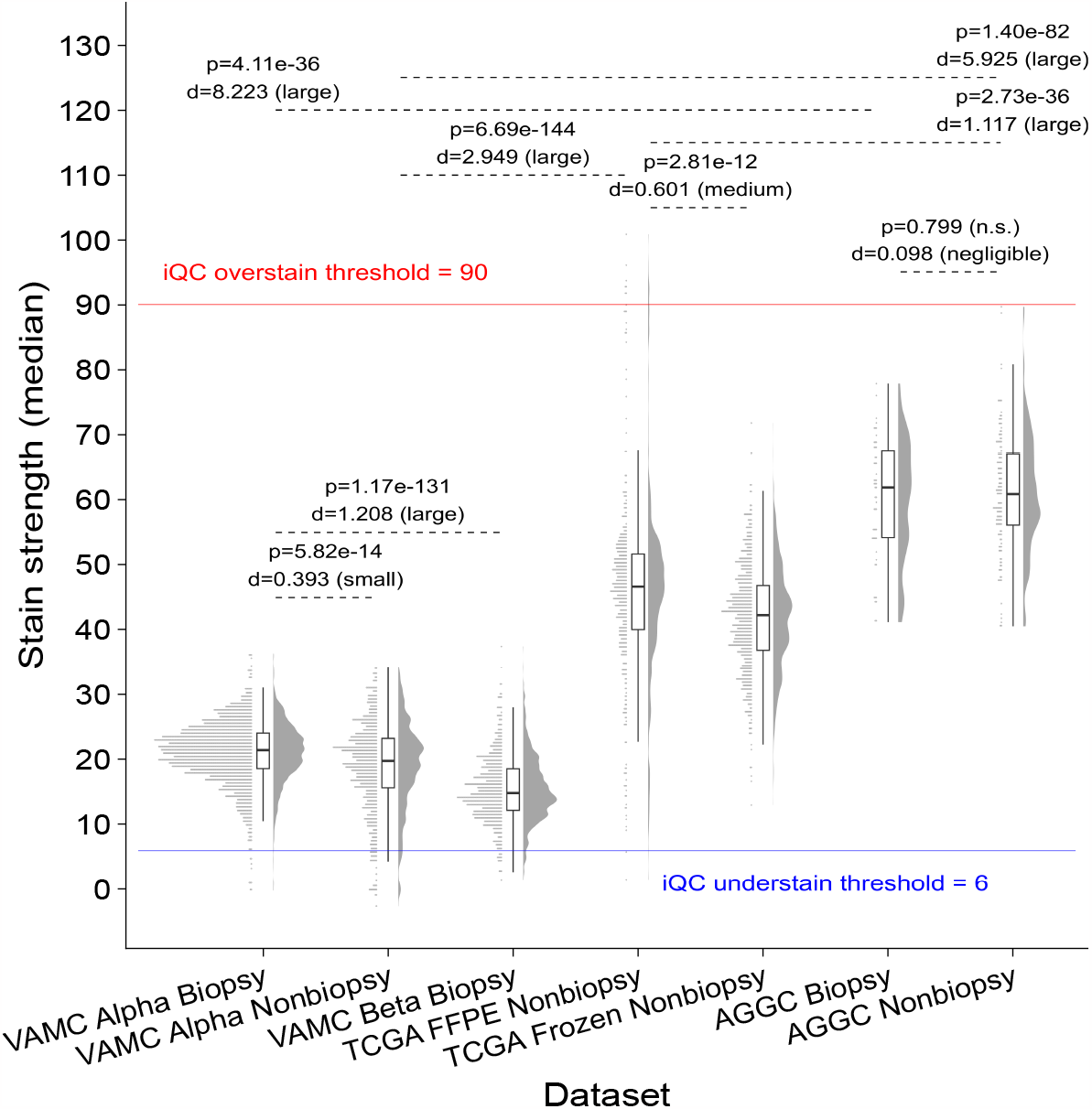
iQC stain strength quantifies differences among VAMCs, datasets, and surgical procedures. For the AGGC dataset (*top right*), there is not a significant difference (*p* = 0.7991, two-tailed Wilcoxon rank-sum test) between biospy and nonbiopsy stain strength, which is a negative control for us. For Institution Alpha (*at left*), there is a statistically significant (*p* = 5.823 *×* 10^*−*14^) but small difference (Cohen’s *d* = 0.3930) in stain strength, when comparing biopsies to nonbiopsies, but we do not believe such a small difference is clinically meaningful. When comparing biopsies from Institution Alpha to Institution Beta (*at center*), we find the stain strength differences are both significant (*p <* 2.2 *×* 10^*−*16^) and large (*d* = 1.208), supporting our qualitative finding that Institution Beta slides have faded stains. There is an even larger stain strength difference (*d* = 8.223, *p <* 2.2 *×* 10^*−*16^) between AGGC biopsies and Institution Alpha biopsies, which may be attributed to differences in staining protocols and whole slide scanners.

#### 2.3.2 Negative control: differences in stain strength between biopsies and nonbiopsies at a VAMC unlikely to be clinically meaningful

We found VAMC Institution *α* biopsy slides were stained significantly stronger than *α* nonbiopsy slides (*p* = 5.823*×*10^*−*14^), but this effect was so small (*d* = 0.393) that we believe this difference is probably not clinically meaningful (Fig 7).

#### 2.3.3 Negative control: no differences in stain strength in AGGC biopsies vs nonbiopsies

We found AGGC biopsies and nonbiopsies have stain strength differences that may be due to change alone (*p* = 0.7991), and the overall effect size of stain strength differences was negligible (*d* = 0.0976) (Fig 7). We believe this is an important negative control for iQC’s stain strength statistic, which is particularly valuable because AGGC is a public dataset.

#### 2.3.4 Positive control: large differences in stain strength between VAMCs and AGGC dataset

Highlighting potential differences in slide staining protocols, whole slide scanners, etc – we found AGGC biopsies were stained significantly stronger than VAMC Institution *α* biopsies (*p* = 4.11*×*10^*−*36^), and this effect was large indeed (*d* = 8.223) (Fig 7). We are encouraged that despite these large differences between VAMC and AGGC datasets, iQC could still accurately distinguish biopsies from nonbiopsies for both VAMC and AGGC datasets (Fig 6), which may be evidence that iQC’s methods will generalize well to yet other high-quality non-VAMC datasets.

### 2.4 iQC’s tissue texture statistic quantifies differences among datasets and surgical procedures

We developed a hypothesis that as slide’s stain fades, the observable visual textures in the slide decrease. To test if iQC could quantify differences in tissue texture among VAMCs, among surgical procedures, and between VAMC and non-VAMC datasets, we considered iQC’s novel tissue texture statistic for each whole slide image (Fig 8).

**Fig 8.**
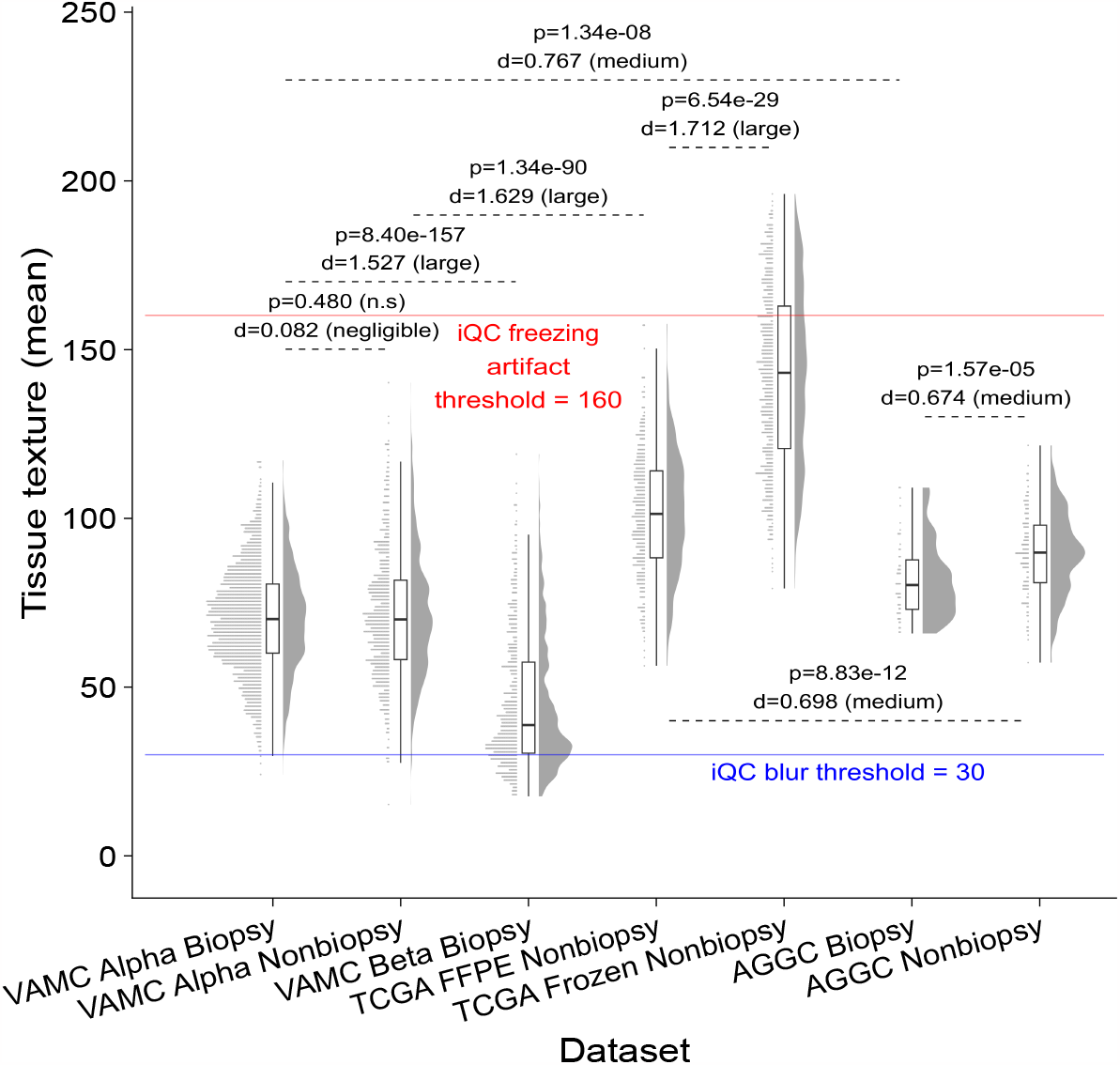
iQC’s nonsuspect tissue texture statistics quantify differences among VAMCs, datasets, and surgical procedures. VAMC Beta biopsies (*third from left on X axis*) have low texture values, because tissue with faded stains do not have high contrast between pixels. TCGA frozen nonbiopsies (*fifth from left on X axis*) have very high tissue texture, because freezing artifacts a.k.a. “swiss cheese artifacts” appear as many small holes in the sample due to small ice crystals breaking apart the tissue. There are a number of textural differences among datasets and procedures, highlighting how different these tissue samples and protocols may be. TCGA may have the widest distribution of texture values because numerous institutions contribute slides to the TCGA consortium.

#### 2.4.1 Significant and large tissue texture difference among VAMC biopsies

We found VAMC Institution *α* contributed biopsy slides that had significantly more visual texture information than VAMC Institution *β*’s biopsy slides (*p* = 8.40*×*10^*−*157^, two-tailed Wilcoxon rank-sum test), and this effect size was large (Cohen’s *d* = 1.527) (Fig 8). This may suggest a slide’s visual texture becomes homogeneous as morphological details fade away with the stain, driving iQC’s tissue texture metric down. This corroborates our pathologist’s expert opinion that *β* slides were faded such that *β* slides were unsuitable for diagnosis (Fig 2F1,G1,H1), and we suspect this large effect as measured by Cohen’s *d* is clinically meaningful.

Indeed, most of the VAMC slides that iQC assigned to the “fail_all_tissue” category (Fig 3A2) were due to tissue texture being below iQC’s blur threshold (Fig 8 blue line).

#### 2.4.2 Increased tissue texture may suggest freezing artifacts

Indicating tissue texture may distinguish slide preparation protocol differences, we find TCGA frozen sections have significantly greater tissue texture than TCGA formalin-fixed paraffin-embedded (FFPE) slides (*p* = 6.54*×*10^*−*29^, two-tailed Wilcoxon rank-sum test) and this effect is large (Cohen’s *d* = 1.712). Freezing makes water ice crystals in the tissue, and these crystals make small rips in the tissue (Fig 5C1,D1). iQC’s tissue texture metric may increase as rips make the tissue adjacent to more of the white background of the slide.

### 2.5 iQC and HistoQC include/exclude statuses for slides are not equivalent

For VAMC and TCGA data, we asked if some configuration of HistoQC may be equivalent to iQC, i.e. may both HistoQC and iQC agree which slides should be included or excluded to a study. If there were such an equivalence between iQC and HistoQC, this may suggest iQC may be less necessary for rigorous quality control if HistoQC were properly configured and used for quality control. Indeed, both iQC and HistoQC uncovered the batch effect from Institution *β* slides (Sec 2.1.1).

#### 2.5.1 iQC and HistoQC agree on VAMC slide exclusion rates in a HistoQC-configuration-dependent manner

As a first step in our equivalency analyses, we asked if iQC may have a comparable slide exclusion rate to some configuration of HistoQC, across all VAMC slides. iQC excludes (“fail_all_tissue” or “fail_some_tissue”, Fig 3A2) 732 of 3673 (19.93%) VAMC slides. In contrast, with its default configuration, HistoQC excludes (“no_tissue_detected”, Fig 3A3) 3336 of 3673 (90.8%) VAMC slides. Rescuing this somewhat with our custom HistoQC configuration file, HistoQC excludes (Fig 3A4) 525 of 3763 (14.29%) VAMC slides, which is significantly less than the number of slides excluded by iQC (*p* = 1.466*×*10^*−*20^, two-tailed binomial test) but this is a negligible effect size (Cohen’s *h* = 0.1501). Therefore, slide exclusion rates differences may not differ to a meaningful degree between iQC and HistoQC with our custom configuration file. We note slide exclusion rate differences do not measure if iQC and HistoQC agree on which specific slides should be excluded.

#### 2.5.2 iQC and HistoQC agree on VAMC slide include/exclude status in a HistoQC-configuration-dependent manner

As a second step in our equivalency analyses, we asked if changing the HistoQC configuration significantly changed HistoQC’s agreement with iQC for VAMC slide include/exclude status. To measure the change in agreement, we considered the odds ratio between iQC and some configuration of HistoQC. We find the odds ratio between iQC and the default configuration of HistoQC was 0.5631 (95% confidence interval (CI) 0.4392-0.7308), while the odds ratio between iQC and our custom configuration of HistoQC was 3.6372 (95% CI 2.9688-4.4529). Because these confidence intervals do not overlap, we conclude our custom configuration for HistoQC significantly improved the slide include/exclude status agreement with iQC, with respect to the default configuration of HistoQC. We find the Matthew’s Correlation Coefficient (MCC) corroborates our use of odds ratios here, specifically that iQC and HistoQC with the default configuration may be anticorrelated (Table 5, MCC = -0.07517, OR = 0.5631), while iQC and HistoQC with our custom configuration may be correlated (Table 5, MCC = 0.2208, OR = 3.6372).

Although we use odds ratio confidence intervals to show the relative agreement significantly changed when reconfiguring HistoQC, we are not aware of widely-accepted thresholds to qualitatively label odds ratios (OR) as “negligible”, “small”, “medium”, or “large” effect sizes. For instance, Chen and colleagues use simulations to compare Cohen’s *d* to OR, to derive thresholds for “negligible” (OR *<* 1.68) “small” (OR *<* 3.47), “medium” (OR *<* 6.71), and “large” (OR≥6.71)[19]. Later, Serdar and colleagues suggested thresholds “negligible” (OR *<* 1.5), “small” (OR *<* 2), “medium” (OR *<* 3), and “large” (OR≥3)[20]. In epidemiology, there has been debate if OR≥3 is too high of a threshold for a clinically meaningful effect[21, 22]. To quote Kraemer and colleagues, “[T]here are no agreed-upon standards for what represents a large odds ratio because some very large odds radios are obtained for situations very close to random association. Consequently, odds ratios can be quite misleading as an effect size indicating clinical significance[23].” Therefore, we consider the phi coefficient (*φ*) and Cohen’s omega (*ω*) as measures of effect size with widely-accepted thresholds for qualitative labels (“negligible”, “small”, etc), below.

#### 2.5.3 iQC and HistoQC agree on VAMC slide include/exclude status to at most a small degree and are not equivalent

As a third step in our equivalency analyses, we asked if iQC slide include/exclude status agreed with HistoQC under various configurations, where the effect size of this agreement could be qualitatively interpreted as “negligible”, “small”, etc according to widely-accepted thresholds. Though we find a statistically significant agreement between iQC and the default configuration of HistoQC (Table 1; *p <* 1.561 10^5^, two-tailed Fisher’s Exact Test), the effect size is negligible (*φ* = 0.07399, Cohen’s *ω* = 0.07517). Additionally, there is significant agreement between iQC and our custom configuration of HistoQC (Table 2; *p <* 2.46*×*10^*−*35^, two-tailed Fisher’s Exact Test), but this is only a small effect size (*φ* = 0.2198, Cohen’s *ω* = 0.2208). Taken together for VAMC slides, this may suggest any agreement between iQC and HistoQC may have at most a small effect size (Table 5). Because HistoQC and iQC share only a small degree of agreement for which VAMC slides should be included or excluded, we conclude HistoQC is not equivalent to iQC for VAMC slides. Effect size thresholds (“negligible”, “small”, etc) are discussed further in Section S1.5.

**Table 1.**
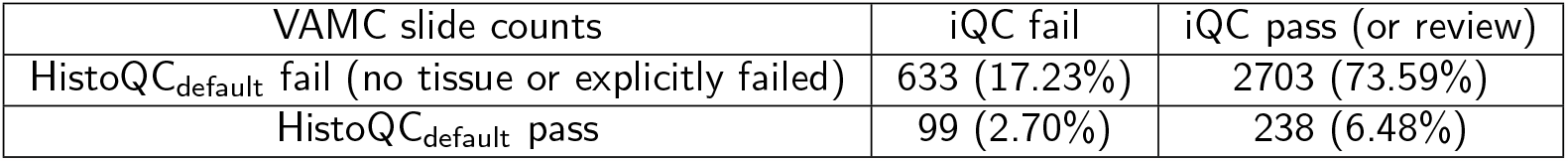
VAMC slide count comparison table for slides that passed/failed iQC (c.f. Fig 3A2) versus passed/failed HistoQC (c.f. Fig 3A3) with HistoQC’s default configuration file. Configured this way, HistoQC and iQC pass most slides. The association between HistoQC and iQC pass/fail statuses for VAMC slides may not be due to chance alone (*p* = 1.561*×*10^*−*5^, two-tailed Fisher’s Exact Test). The odds ratio is 0.5631 (95% CI 0.4392-0.7308), Matthew’s Correlation Coefficient is -0.07517, phi coefficient is 0.07399, and Cohen’s *ω* is 0.07517 indicating a negligible degree of association between the iQC and HistoQC pass/fail statuses of VAMC slides.

**Table 2.** VAMC slide count comparison table for slides that passed/failed iQC (c.f. Fig 3A2) versus passed/failed HistoQC (c.f. Fig 3A4) with our custom HistoQC configuration file. Configured this way, HistoQC and iQC pass most slides. The association between HistoQC and iQC pass/fail statuses for VAMC slides may not be due to chance alone (*p* = 2.46*×*10^*−*35^, two-tailed Fisher’s Exact Test). The odds ratio is 3.6372 (95% CI 2.9688-4.4529), Matthew’s Correlation Coefficient is 0.2208, phi coefficient is 0.2198, and Cohen’s *ω* is 0.2208 indicating only a small degree of association between the iQC and HistoQC pass/fail statuses of VAMC slides.

| VAMC slide counts | iQC fail | iQC pass (or review) |
| --- | --- | --- |
| HistoQC<sub>custom</sub> fail (no tissue or explicitly failed) | 218 (5.94%) | 307 (8.36%) |
| HistoQC<sub>custom</sub> pass | 514 (13.99%) | 2634 (71.71%) |

#### 2.5.4 iQC and HistoQC agreement on TCGA slides may be due to chance alone and are not equivalent

iQC excludes (“fail_all_tissue” or “fail some tissue”, Fig 3A7) 7 of 1172 (0.597%) TCGA slides. With its default configuration, HistoQC excludes (“no_tissue_detected”, Fig 3A5) 70 of 1172 (5.97%) TCGA slides. With our custom HistoQC configuration file, HistoQC excludes (Fig 3A6) zero of 1172 (0%) TCGA slides, which is significantly less than the number of slides excluded by iQC (*p* = 0.001813, two-tailed binomial test) but this is a negligible effect (Cohen’s *h* = 0.1547), so exclusion rates may not differ to a meaningful degree between iQC and HistoQC with our custom configuration file. For completeness, we compare iQC to both configurations of HistoQC, below.

We do not find a statistically significant agreement between iQC and the default configuration of HistoQC (Table 3, *p* = 0.3509), and the size of this effect is negligible (*φ* = 0.2198, *ω* = 0.02719). Additionally, there is not a statistically significant agreement between iQC and our custom configuration of HistoQC (Table 4, *p* = 1). Taken together for TCGA slides, this may suggest any agreement between iQC and HistoQC may be due to chance alone. Therefore, we conclude HistoQC is not equivalent to iQC for TCGA slides.

**Table 3.**
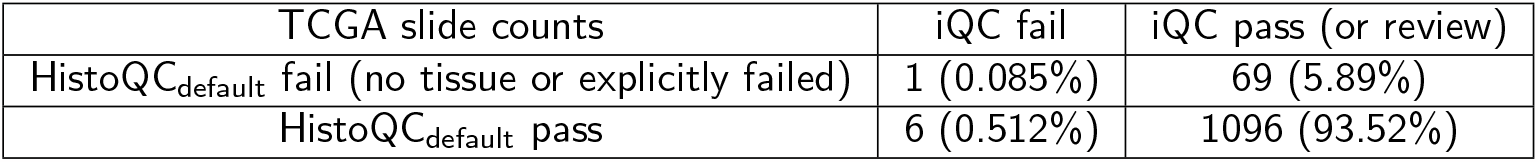
TCGA slide count comparison table for slides that iQC included/excluded (c.f. Fig 3A7) versus HistoQC included/excluded (c.f. Fig 3A5), with HistoQC’s default configuration file. Configured this way, HistoQC and iQC include most slides. The association between HistoQC and iQC include/exclude statuses for TCGA slides may be due to chance alone (*p* = 0.3509, two-tailed Fisher’s Exact Test). The odds ratio is 2.6429 (95% CI 0.05674-22.26), Matthew’s Correlation Coefficient is 0.02719, phi coefficient is 0.003828, and Cohen’s *ω* is 0.02719. Taken together, the association between the iQC and HistoQC include/exclude statuses of TCGA slides may be due to chance alone, and the size of any association effect is negligible, so we would not expect the association to be clinically meaningful.

**Table 4.** TCGA slide count comparison table for slides that iQC included/excluded (c.f. Fig 3A7) versus HistoQC included/excluded (c.f. Fig 3A6), with our custom configuration file for HistoQC. Configured this way, HistoQC includes all slides and iQC includes all but seven. The association between HistoQC and iQC include/exclude statuses for TCGA slides may be due to chance alone (*p* = 1, two-tailed Fisher’s Exact Test). Because there are zero slides that iQC and HistoQC both exclude, the odds ratio, Matthew’s Correlation Coefficient, phi coefficient, and Cohen’s *ω* are undefined. Taken together, the evidence does not suggest an association between the iQC and HistoQC include/exclude statuses of TCGA slides.

| TCGA slide counts | iQC fail | iQC pass (or review) |
| --- | --- | --- |
| HistoQC<sub>custom</sub> fail (no tissue or explicitly failed) | 0 (0%) | 0 (0%) |
| HistoQC<sub>custom</sub> pass | 7 (0.597%) | 1165 (99.40%) |

**Table 5.**
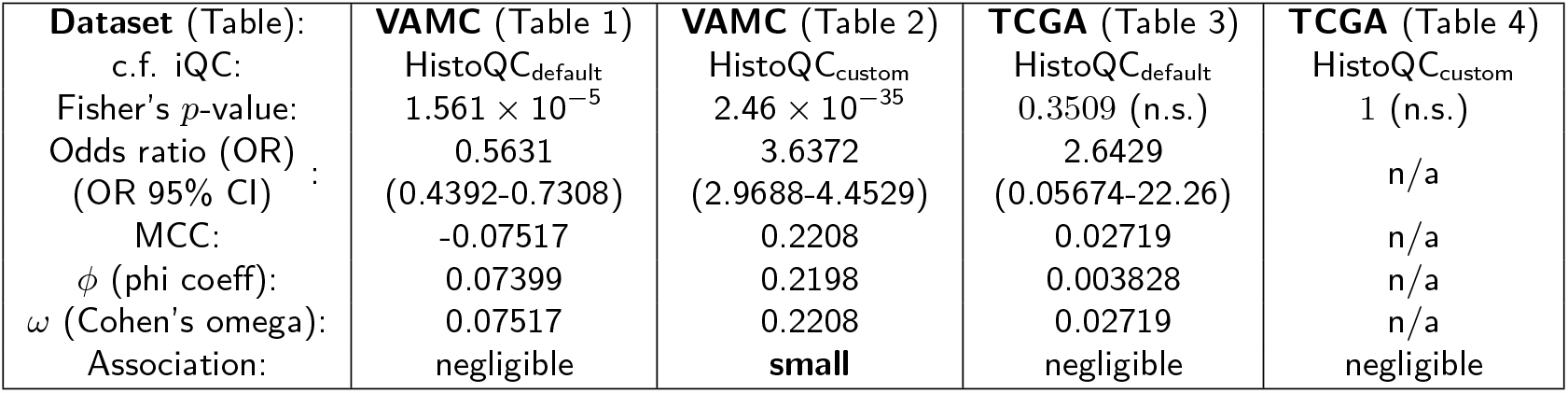
Non-equivalency of iQC and HistoQC across VAMC and TCGA datasets. For VAMC, if iQC include/exclude statuses for slides are compared to HistoQC with its default configuration, there is a statistically significant p-value (1.561*×*10^*−*5^) that may suggest the association between iQC and HistoQC pass/fail statuses for slides may not be due to chance alone, but the size of this effect is negligible (*φ* = 0.1101), so we would not expect this association to be clinically meaningful. Moreover, for TCGA, the iQC and HistoQC association is both negligible and not statistically significant. Only for VAMC data where iQC is compared to HistoQC with our custom configuration do we observe a small degree of association between iQC and HistoQC. Taken together, we believe this evidence may suggest iQC and HistoQC include/exclude statuses for slides across these datasets are not equivalent.

| Dataset (Table): | VAMC (Table 1) | VAMC (Table 2) | TCGA (Table 3) | TCGA (Table 4) |
| --- | --- | --- | --- | --- |
| c.f. iQC: | HistoQC<sub>default</sub> | HistoQC<sub>custom</sub> | HistoQC<sub>default</sub> | HistoQC<sub>custom</sub> |
| Fisher's $p$-value: | $1.561 \times 10^{-5}$ | $2.46 \times 10^{-35}$ | 0.3509 (n.s.) | 1 (n.s.) |
| Odds ratio (OR) : | 0.5631 | 3.6372 | 2.6429 | n/a |
| (OR 95% CI) : | (0.4392-0.7308) | (2.9688-4.4529) | (0.05674-22.26) | n/a |
| MCC: | -0.07517 | 0.2208 | 0.02719 | n/a |
| $\phi$ (phi coeff): | 0.07399 | 0.2198 | 0.003828 | n/a |
| $\omega$ (Cohen's omega): | 0.07517 | 0.2208 | 0.02719 | n/a |
| Association: | negligible | **small** | negligible | negligible |

We summarize our equivalency study in Table 5 and conclude iQC is not equivalent to HistoQC in slide include/exclude statuses, for the two HistoQC configurations we tested, across VAMC and TCGA datasets. It appears HistoQC largely excludes slides when tissue is not detected, which may be a different failure mode than iQC excluding slides due to faded stain or age-related bubbles (Figs 3, 4, and 5). Because of the nonequivalency of iQC and HistoQC include/exclude statuses, we do not perform a more detailed quantitative per-pixel equivalency study of iQC and HistoQC masks, although we do compare masks qualitatively (Figs 4 and 5).

## 3 Methods

This study was approved by the Institutional Review Board at the VA Boston Healthcare System.

Institutions *α* and *γ* prepare slides with a special Toluene-based mounting media (Fig 1B2a) that is resistant to oxidation, discoloration, and fading (Sec S1.6). This may contribute to the quality of slides from Institutions *α* and *γ*. We suspect Institution *β* used mounting tape (Fig 1B2b) to prepare many or all of their slides.

iQC generates interpretable statistics to calculate a score for biopsy/nonbiopsy prediction. iQC has a multistep pipeline, including (1) Otsu thresholds (Sec S1.2.1), (2) preliminary pixel typing (Sec S1.2.2), (3) debris detection (Sec S1.2.3), (4) edge detection (Sec S1.2.4), (5) blur artifact detection (Sec S1.2.5), (6) black mark detection (Sec S1.2.6), (7) pen detection, connection, and extension (Sec S1.2.7), (8) stain strength calculations (Sec S1.2.7.1), (9) write 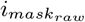mask (Sec S1.2.8), (10) blur artifact orientation detection (Sec S1.2.9), (11) writing of 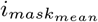 that mixes base image with 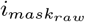 (Sec S1.2.10), (12) machine-learning-driven inference of “suspect” type pixels to other types e.g. pen, tissue, and background (Sec S1.2.11), (13) write 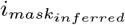 mask (Sec S1.2.12), (14) biopsy/nonbiopsy prediction (Sec S1.2.13), (15) ridge detection, age-related bubble detection (Sec S1.2.14), and “window pane breaking” detection (Fig S1), (16) write 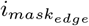 mask (Sec S1.2.15), (17) barcode and text detection for PHI/PII risks (Sec S1.2.16), and (18) close-out timing statistics (Sec S1.2.17).

iQC’s biopsy/nonbiopsy predictor is a function that generates a score 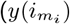 in Eqn 1) between 0 and 1000000, with low numbers favoring a biopsy and high numbers favoring a nonbiopsy, e.g. prostatectomy or TURP, where for brevity we denote 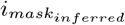 as 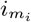:

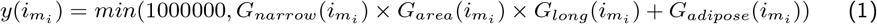

Each *G*(…) is a Gompertz function[24], further discussed in the supplement (Sec S1.4).

Computational software and hardware (Sec S1.5), as well as whole slide image details (Sec S1.7), are detailed in the supplement.

## 4 Discussion

### 4.1 Data quality defined on objective ground truth

To our knowledge, we are the first to define quality in terms of objective ground truth data, e.g. a dataset is high quality if the AUROC is close to 1.0 for a surgical procedure prediction task. Surgical procedure data are available from a Laboratory Information Management System (LIMS, Fig 1D1b). For iQC, AUROC is close to 1.0 for all datasets and VAMCs except Institution *β*. Institution *β* has AUROC of 0.71 and poor quality data (Fig 6C).

#### 4.1.1 Objective ground truth from LIMS or EHR is scalable

We believe iQC is a novel approach to quality control because it is rooted in objective ground truth. Highly curated surgical procedure or other coded data from the LIMS or Electronic Health Record (EHR) are readily available. This approach may scale well because surgical procedure annotations are at the whole slide level, rather than at the region of interest (ROI) or pixel level. Slide-level annotations drive the scalability of Campanella[14], Lu[15], and other weakly supervised learning pipelines. Still, iQC provides quality control masks for per-pixel semantic segmentation, e.g. 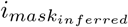, to assist pathologists and downstream AI pipelines in distinguishing artifacts such as pen (Fig 5E2) or blur (Fig 2A3,B3) from tissue in the whole slide image.

#### 4.1.2 Prior work in defining quality

iQC’s definition of quality in terms of objective ground truth data differs from some prior approaches to quality control in digital pathology. In 2019, Schaumberg and Fuchs defined quality control in terms of numeric parameters or rules that were hand-engineered[7]. To refine the subjective nature of quality control, HistoQC validated quality control in terms of pathologist concordance[25]. A follow-up HistoQC study of the Nephrotic Syndrome Study Network (NEPTUNE) digital pathology repository disqualified 9% of slides as unsuitable for analysis, where slides were disqualified if particular statistics were out of pre-programmed bounds, such as the mean brightness of the entire slide being too high or low[26]. HistoROI claimed to improve upon HistoQC through human-in-the-loop training and a deep learning system on annotated tiles, although this quality control still depends on manual annotations from pathologists and their subjective interpretation of the morphology, which may be challenging to independently reproduce exactly[27].

### 4.2 Myriad technical factors drive quality

Institution *β* slides were made in the years 2000-2007. The slides were over a decade old when they were scanned in 2023 (Sec S1.7). The slides from the other VAMCs were made in the years 2003-2021. We believe this suggests there are myriad technical factors beyond calendar age that contribute to how over time slides show signs of age, e.g. faded stain (Sec S1.2.7.1) and acrylamide bubbles (Sec S1.2.14). Technical factors may include the formulation of hematoxylin and eosin (H&E) stains, the slide mounting protocol (e.g. Fig 1B2a mounting solution vs Fig 1B2b mounting tape), and storage conditions of the slides (Fig 1B3). Storage conditions may vary in terms of temperature, humidity, or ultraviolet light exposure. Institution *α*’s protocols may have mitigated some of these technical factors to maintain high quality (Sec S1.6).

### 4.3 Pathologist review essential for assessing quality, taking action

iQC detected the faded stain and assigned many Institution *β* slides to the “fail_all_slide” quality control category to indicate these slides are not suitable for diagnosis (Fig 3A2). An anatomic pathologist later reviewed the slides and recommended all Institution *β* slides be (i) excluded from studies and (ii) physically remade (e.g. Fig 3B2). iQC’s stain strength statistic recapitulated the pathologist’s finding of faded stains at Institution *β* (Fig 7), as did iQC’s tissue texture statistic (Fig 8).

### 4.4 Quality control machine learning framed as search

Proceeding from some of our earlier work that showed how tractable search is in computational pathology[28, 29], we framed quality control as a search problem at the pixel level. Rather than train artifact-specific classifiers to detect blur[3], pen[4], or coverslip breaks[6], iQC uses machine learning to compute how similar a “suspect” pixel is to other “nonsuspect” pixel types (e.g. pen, tissue, background, etc see Section S1.8). We believe this allows iQC to define relatively simple criteria for the appearance of different pixel types, and extend these rules using machine-learning-driven inferences (Fig 2P1-P3), to achieve high AUROC performance across VA and AGGC datasets (Fig 6).

## 5 Conclusion

Our iQC pipeline found a batch effect from a medical center that provided aged poor-quality slides to scan, where the inexpensive preparation of the slides may have interacted with the adverse slide storage conditions at the medical center. Moreover, iQC provides type information for each pixel, uses pixel type information to predict surgical procedure, and provides an overall AUROC for surgical procedure prediction performance that may be positively associated to the overall quality of data produced at a medical center. iQC provides numerous statistics to describe each slide, including novel stain strength and tissue texture statistics that each corroborate the pathologist’s finding that slides from this specific medical center were unsuitable for rendering a diagnosis. iQC reports provide quality feedback to a medical center, e.g. to restain specific slides.

We find high AUROC for all datasets and medical centers except the one medical center with aged slides. At this medical center (*β*) AUROC is correspondingly much lower and histopathology stains are faded. Because iQC separates biopsies from nonbiopsies in mixed incoming datasets, we believe iQC may be especially valuable for downstream studies where only biopsies may be included in a study, to the exclusion of all other surgical procedures, i.e. prostatectomies, TURPs, colonic polypectomies, etc.

One configuration and implementation of iQC appears to generalize well across VAMC, AGGC, and TCGA datasets – insofar as iQC reliably detects tissue from each of these data sources. This is in contrast to HistoQC’s default configuration, which reliably detects tissue in TCGA but excludes most VAMC slides as “no_tissue_detected”. Our custom configuration file for HistoQC did reliably detect tissue in both VAMC and TCGA data, but this configuration may exclude some adipose tissue and retain some artifacts. Across VAMC and TCGA datasets, for both HistoQC’s default configuration and our custom HistoQC configuration, we found a negligible to small degree of slide exclusion status agreement between iQC and HistoQC, which may suggest iQC and HistoQC are not equivalent in how they include or exclude slides.

To our knowledge, we present the first quality control pipeline for histopathology validated to objective ground truth evidence, specifically surgical procedure. Following this approach, a hospital may apply our quality control pipeline and validate against surgical procedure data in their LIMS or EHR, without requiring effort or annotations from pathologists. We encourage broad adoption of such scalable quality control pipelines in digital pathology and computational pathology pipelines.

## Data Availability

All data produced in the present study are available in a de-identified format on the Veterans Affairs GenISIS platform for researchers upon request and approval.

## 6 Acknowledgements

This work was funded through a Prostate Cancer Foundation grant PCFCHAL22 to MBR, BSK, IPG, and SP. Support for the GenISIS datacenter was additionally provided by the United States Veterans Administration (VA) Office of Research and Development (ORD) through SP. Authors thank Mark Hewitt and Nicholas Burns for high performance computing support. AJS thanks Dr Mariam Aly for early manuscript discussion, for The Noun Project recommendation (Sec S1.9), and for Cohen’s *d* analysis recommendations. We are grateful to the patients who made this study possible. The content is solely the responsibility of the authors and does not necessarily represent the official views of the U.S. Department of Veterans Affairs, the Department of Defense, or the United States Government.

## 7 Contributions

Conceptualization: AJS, SP.

Data acquisition: AJS and RK (AGGC and TCGA slides), AW and NK (VAMC slides),

NW (VAMC LIMS biopsy/nonbiopsy surgical procedure metadata).

Data curation: AJS, MSL.

Data transfer and management: AJS, RN, AW, NK, GT, PK, PD, NW, RK.

Methodology, software, validation, formal analysis, investigation, visualization, writing

(original draft): AJS.

Funding acquisition: MBR, BSK, IPG, SP.

Project administration: AJS, MSL, RN, AW, NK, MBR, BSK, IGP, SP.

Resources (pathology) and discussion: MSL, RN, AW, NK, BSK, IPG.

Resources (computational) and discussion: GT, PK, PD, RK, SP.

Supervision (pathology): MSL, RN, IPG.

Supervision (data transfer): RN, RK, IPG, SP.

Supervision (computational): MSL, SP. Writing (editing): AJS, RN.

Writing (reviewing): AJS, RN, PK, BSK, IPG, SP.

## 8 Ethics Declaration and Conflicts of Interest

The author(s) declare they have no competing interests.

## Supporting Information

### S1 Supplementary materials and methods

#### S1.1 iQC quality control categories

iQC defines ten quality control categories (Fig 3). This provides granular information for the quality of a slide. These categories are grouped into “fail…”, “review…”, and “pass…” supercategories. We define each of the ten categories below.

1. fail_all_tissue: iQC suggests all tissue in the slide is not suitable for any diagnostic purpose. The tissue is not suitable for a pathologist to render a diagnosis. The tissue is also not suitable for downstream computational analysis / machine learning / artificial intelligence (AI). Typically, this occurs because the tissue staining is badly faded (Sec S1.2.7.1), e.g. the hematoxylin stain is not visible and only eosin remains. This fading worsens as the slide ages, depending on the storage conditions, the quality of stains used, and perhaps other factors (Sec S1.6). Equations S4 and S8 define iQC’s metrics to determine “fail_all_tissue” status.
2. fail_some_tissue: iQC suggests some of the tissue is not suitable for any diagnostic purpose. This typically occurs if the mounting solution/layer that adheres the glass coverslip to the glass slide has aged (Sec S1.6). The mounting solution may break down to form acrylamide, leading to “window pane breaking” artifacts (Fig S1) and bubbles. If these artifacts or bubbles occur over tissue in the slide, such occluded tissue may not be suitable for a pathologist or AI. These regions should be excluded, while the rest of the unaffected tissue may be retained and used by a pathologist or AI. It may be especially problematic if such artifacts or bubbles occlude all the malignant foci in the slide, or other foci of disease. Such occlusion may change the diagnosis, depending on whether or not disease foci are included or excluded. For this reason, great care should be taken when using fail_some_tissue slides. Equation S9 defines iQC’s metrics to determine “fail_some_tissue” status.
3. review_for_inadequate_tissue: iQC suggests there is very little tissue in the slide. This slide should be manually reviewed by an expert to determine if sufficient tissue exists for either a pathologist to diagnose a disease or an AI to analyze.
4. review_for_acrylamide_aging: iQC suggests there may be evidence of acrylamide aging in the slide, e.g. window pane breaking artifacts or bubbles. The evidence is not strong, so manual expert review is recommended.
5. review_for_systematic_blur: iQC suggests there may be evidence of a specific type of blur in the slide, which we call systematic blur. Systematic blur is thought to occur when the acrylamide layer has aged such that the glass coverslip is not securely adhered to the glass slide, so the slide “shakes in place” while the slide is being scanned, and this shaking is such that the scanner’s autofocus cannot focus correctly on the slide to get a sharp picture. The result is a band of blurred pixels, e.g. a horizontal band of blurring as the scanner’s camera travels left to right to photograph parts of the slide. Systematic blur induces subtle linear artifacts in the background between adjacent passes of the scanner’s camera, which may occur when the scanner’s software stitches together images. iQC detects these linear artifacts as straight lines of “suspect” pixels. If the number of such linear suspect pixels exceeds a threshold, iQC recommends the slide for manual expert review here. This may be a novel way to detect blur, in that we look at lines in the slide background, rather than directly look for blurry pixels.
6. review_for_faded_stain: iQC suggests there may be evidence of stain fading in the slide, but this evidence is not strong, so manual expert review is recommended.
7. review_for_debris_pen_etc: iQC suggests there may be evidence for debris, pen, or marker in the slide. Manual expert review is recommended. Some care should be taken with these slides, e.g. if pen marks occlude foci of disease, omitting such foci may change the diagnosis.
8. review_for_barcodes_writing_pii_etc: iQC introduces a potentially novel method to detect barcodes or other black structured marks (like text) on a slide. Slides with names printed on them in black text are not de-identified and are not suitable for research purposes. Often, however, the printed text in a slide indicates where the slide was manufactured, rather than indicating PHI/PII of the patient. Manual expert review is recommended.
9. pass_intact: iQC suggests the slide is generally good condition, though there may be some small amount of pen, marker, or debris present in the slide. Automated tools such as iQC may recommend where pen, marker, etc are in the slide so these may be avoided. iQC recommends the slide is otherwise of high quality and is expected to be suitable for both a pathologist and AI.
10. pass_pristine: iQC suggests the entire slide is high quality and can likely be used as-is.

#### S1.2 iQC algorithm steps

iQC has a number of steps outlined below. iQC operates on the whole slide image at 10x total magnification and expects a scan at 0.25 microns per pixel. Resolution is adjusted to compensate if the scan is 0.5 microns per pixel.

##### S1.2.1 Otsu threholds

iQC calculates thresholds via Otsu’s Method[13]. Thresholds are calculated for the red channel, green channel, blue channel, grayscale pixel values, and Sobel magnitude values. For machine learning, a pixel is represented by a 4-dimensional red, green, blue, and Sobel vector of values (Sec S1.8).

##### S1.2.2 Preliminary pixel typing

iQC uses simple rules to assign a preliminary type to each pixel, where the types are hematoxylin type, red type, green type, blue type, background type, black type, and tissue type. The red/green/blue/black types are often pen or marker. The red pixel type may alternatively represent blood/erythrocytes, which may be over-represented in slides overstained with eosin. The black types may also be debris, tissue folds, or necrosis. The hematoxylin type is a special subtype of the tissue type. Hematoxylin types have a blue channel value greater than red channel value, a red greater than green, and a Sobel value above the corresponding Otsu threshold (Sec S1.2.1).

##### S1.2.3 Debris detection

If a set of nonbackground pixels are completely enclosed in box with sides of length 2*r*, then the enclosed nonbackground pixels are typed as debris. iQC lets *r* = 10, e.g. the debris box radius is 10. The 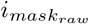 and 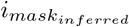 indicate debris pixels in brown (e.g. Figs 4E1, 5B2,C2 and 6F2).

##### S1.2.4 Edge detection

When slide mounting solution or mounting tape (Fig 1B2a,b) ages, “window pane breaking” artifacts and acrylamide bubbles may form. Both these signs have well-defined edges. iQC detects edges to both describe the whole slide image and estimate the presence of these signs of age. iQC excludes debris from edge detection. By excluding debris, the count of edge pixels may be more accurate, so by extension the statistics from the calculation of how many pixels are edges related to window pane breaking or bubbles may be more accurate. iQC uses of Otsu’s Method to type a pixel as an edge or not.

##### S1.2.5 Systematic blur detection via bar artifacts

Horizontal bars of suspect pixels may indicate systematic blur (Fig 2A3,B3). In principle, there could be vertical bars as well, depending on how the scanner scans, either left to right (horizontal) or top to bottom (vertical).

##### S1.2.6 Black mark detection

iQC next converts “suspect” type pixels to “black” type pixels according to grayscale Otsu Thresholds.

##### S1.2.7 Pen detection, connection, and extension

For every red, green, blue, or black “pen” type pixel – iQC attempts to make straight lines connecting two pen pixels having the same type (e.g. red pixel may connect to a red pixel, blue pixel may connect to a blue pixel, etc), then flood fill through suspect pixels that are not stitching artifacts. The extent of flood fill is limited to a specific distance away from where the flood fill started, which prevents flood fill from overwriting large portions of 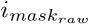 with pen pixel types.

iQC counts tissue and suspect pixels, replacing tissue pixels with pen if tissue pixel is surrounded by pen or background. This is intended to remove any false tissue type pixel perimeter from pen marks.

###### S1.2.7.1 Stain strength calculation

iQC calculates stain strength in two ways, mean stain strength (Eqn S4) and median stain strength (Eqn S8). If most of the tissue is stroma, then the mean or median pixel is expected to approximate the red, green, and blue pixel values of stroma in the slide. If *stain_strength*_*mean*_ is less than *stain_strength*_*mean_threshold*_, or *stain_strength*_*median*_ is less than *stain_strength*_*median_threshold*_, iQC categories the slide as “fail_all_tissue” due to faded stain. Presently, *stain_strength*_*mean_threshold*_ = 6 and *stain_strength*_*median_threshold*_ = 6 (Fig 7 blue line).

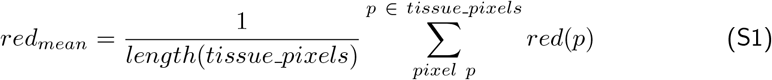

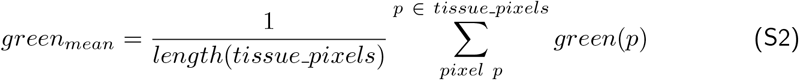

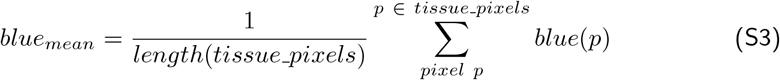

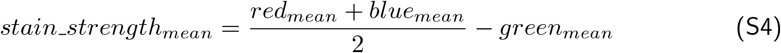

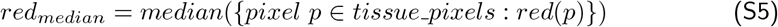

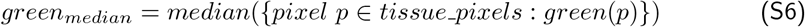

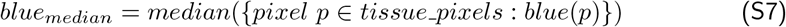

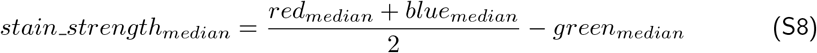

For *median*(…) we use the grouped median rather than simple median, because there are so many duplicate values in a channel for an image, e.g. *red*(*p*) values are integers between 0 and 255 inclusive. We found this median approach to be better behaved than the mean, e.g. via the median there is no significant difference in stain strength between AGGC biopsies and AGGC nonbiopsies, which we believe is an important negative control (Fig 7).

###### S1.2.7.2 Systematic blur threshold

iQC calculates how sharp (i.e. not blurry) the tissue pixels are. This sharpness is defined by a Sobel Magnitude. This sharpness should not be confused with systematic blur. Tissue is visually sharp when tissue pixels tend to have different grayscale values when compared to adjacent pixels. A freshly-stained slide will tend to be more vibrantly-stained and more “sharp” than an aged slide, as fresh stains will highlight differences among tissues well. For instance, it appears Institution *β* slides had both faded stain and measurably less visual texture (Fig 8).

iQC estimates a slide may have evidence of systematic blur if the total number of suspect pixels that participate in horizontal or vertical bars exceeds double the height or width of the slide image at 10x (Fig 2A3,B3). This may be used as a warning to that the slide may benefit from manual review to check for systematic blur.

###### S1.2.7.3 Slide age estimates

iQC estimates slide age as a stain strength metric multiplied by a tissue sharpness metric. The intuition for this is aged slides with faded stain will have both (1) low stain strength because stain has faded to gray, and (2) low tissue sharpness because old stain does not vibrantly highlight differences among adjacent tissues. The intuition continues that for new freshly-stained slides there will have both (1) high stain strength because reds (from eosin) and blues (from hematoxylin) will be much stronger then greens (neither hematoxyin or eosin is green) in the slide and (2) high tissue sharpness because fresh stain will vibrantly highlight differences among adjacent tissues.

##### S1.2.8 Write 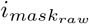

iQC writes 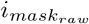 as a semantic segmentation of pixel types in the whole slide image.

##### S1.2.9 Stitching score for systematic blur bar artifacts

iQC detects if stitching artifacts run left-right (horizontally) or up-down (vertically). The blur boundary will be parallel to stitching artifacts, if there is a blur boundary. For example, if there are horizontal stitching artifacts, there will also be horizontal bands of systematic blur.

##### S1.2.10 Write 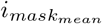

iQC writes a mean image that is a mix of the pixel types with the original pixel values in the slide image.

##### S1.2.11 KNN inference of suspect pixels to other types

Not all pixels fit within iQC’s rigid set of rules for background, pen, tissue, etc (per Sec S1.2.2). Many pixels may be typed as “suspect” in the slide to indicate these pixels may be tissue, but the pixels may also be background, pen, etc. To infer the type of suspect pixels, iQC uses the K-Nearest Neighbors (KNN) machine learning algorithm for pixels (Sec S1.8). Specifically, for a given suspect pixel *p*, iQC uses KNN to find the pixel *c*, where *c* has the most similar red, green, blue, and Sobel magnitude values to *p*. This is sometimes referred to as the “closest pair of points problem”. Then iQC assigns the 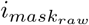 type of *c* to *p*. Thus *p* is no longer the suspect type. Instead, the type of *p* is equal to the type of *c*. This is the inductive bias of KNN, e.g. that the type of a pixel is mostly likely the same as the type of the most similar pixel. This is a very simple inductive bias that is readily interpretable. KNN may be considered the simplest possible machine learning algorithm, so KNN is a logical first choice of machine learning algorithms, by Occam’s Razor. The performance and relative complexity of more advanced machine learning algorithms may then be compared to KNN.

###### S1.2.11.1 Accelerated KNN

Unfortunately, KNN is notoriously slow. Big O notation for the computational complexity of KNN is *O*(*KNN*) *∈ O*(*n*^2^) where *n* is the number of pixels in 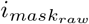 (for brevity, we denote 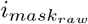 as 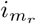). More specifically, if we let 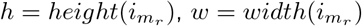, and *s* = (*the total number of suspect pixels in* 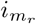), then *O*(*KNN*) *∈ O*(*s × h × w*). This means if there are 1000 suspect pixels to infer, and 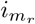 is 1500 pixels high by 3000 pixels wide, then KNN must make 4500000000 (4.5 billion) pixel-to-pixel comparisons. 4500000000 is an extraordinarily high number of computations and is computationally intractable when considering a dataset may easily contain a few thousand whole slide images, e.g. (4500000000 computations/image) *×*(4000 images/dataset)= 18000000000000 (18 quadrillion) computations for a dataset. There are scalability challenges to overcome.

Inspired by our prior work[29], iQC implements speeds up KNN using a hashtable (ak.a. dictionary). For every *pixel*_*jpg*_ in *i*_*jpg*_ (where *i*_*jpg*_ is standardized by iQC to 10x magnification from a 0.25 microns per pixel scan), iQC makes a hashtable “key” from the *pixel*_*jpg*_ red, green, blue, and Sobel magnitude values (a.k.a. RGBS). If there is a suspect pixel with this RGBS value, then this iQC assigns to that suspect pixel the type of the corresponding nonsuspect pixel with the same RGBS value. Importantly, a hashtable lookup is practically instantaneous, i.e. *O*(*hashtable_lookup*) ∈ *O*(1). Loosely speaking, a hashtable lookup may be understood to take one computation, for the purposes of Big O notation. A hashtable lookup is very fast. This approach scales well, e.g. if every suspect pixel may be found in the hashtable, best case performance of KNN is *O*(*KNN_with_hashtable*) ∈ *O*(*s*), or more generally *O*(*KNN_with_hashtable*) ∈ *O*(*n*). This is much more computationally tractable, e.g. if there are on average 1000 suspect pixels to infer per image, the hashtable-accelerated KNN performs 1000 computations on this image, which several orders of magnitude less than 4500000000. We find this approach to be scalable to thousands of images in a dataset, e.g. (1000 computations/image) *×* (4000 images/dataset)= 4000000 (4 million) computations for a dataset. Crucially, most of the time the hashtable must find a pixel with the same RGBS value as a suspect pixel, but this cannot always be guaranteed.

To increase the chance that the hashtable will find a matching RGBS value, iQC performs several hashtable lookups with slightly different RGBS values, e.g. with red incremented or decremented by 1. Hashtable lookups remain fast, and several hashtable lookups is still a scalable way to compute KNN across all pixels. If we let *H* = (*the total number of hashtable lookups to perform for each suspect pixel*), then *O*(*KNN_with_hashtable_and_many_lookups*) ∈ *O*(*s × H*), or more generally *O*(*KNN_with_hashtable_and_many_lookups*) ∈*O*(*n*). That is, adding more hashtable lookups adds a linear computational complexity to KNN, which factors out of the Big O notation. The additional computational complexity is minimal. The choice of *H* may vary depending on the type to infer, e.g. iQC uses a high *H* to infer suspect pixels as background and a much lower *H* to infer pixels as tissue, because there may be some variance in the background color within the slide (due to lighting, artifacts, etc), whereas tissue staining is relatively uniform within a slide. For the majority of the images in our data, we find this hashtable approach finds a KNN match for most pixels in *i*_*jpg*_. If a KNN match is not found through the hashtable, iQC’s algorithm falls back on a naïve KNN, which in practice we find is performed for a minority of pixels, if any, but this can be dataset-dependent. iQC’s approach thus remains computationally tractable and scalable in practice. We find a purely naïve KNN on a dataset does not complete after weeks of computation.

iQC’s hashtable-accelerated KNN approach may be thought of as a “fixed-radius near neighbors” algorithm, in that the red, green, blue, and Sobel magnitude values all fall on an integer lattice, which is used to perform hashtable lookups to find near neighbors. These methods were originally studied in Bentley 1975[30] “A survey of techniques for fixed-radius near neighbor searching”, which cited Levinthal 1966[31], for the purposes of visualizing molecular structures. Levinthal’s task was to consider the interactions involving a specific atom to (a) all other atoms in the atom’s spatial cube and (b) all other atoms in 26 surrounding cubes – which is much less computationally intense than considering all pairwise interactions among all atoms in a protein. Bentley’s task was similarly to quickly identify atoms in a spatial neighborhood, to focus visualization on these atoms. Bentley correctly recognized this approach may be extended to an arbitrary metric space, but he did not elaborate. Our application of fixed radius near neighbors differs from Levinthal’s and Bentley’s tasks in that:

1. iQC’s use of a hashtable for fixed-radius search is in a feature space (i.e. red, green, blue, and Sobel magnitude features of a pixel) rather than a Cartesian coordinate space.
2. iQC’s general use of fixed-radius search is for the purpose of inferring the “type” of pixels in whole slide images, rather than restricting a spatial neighborhood for intermolecular force calculation or submolecular visualization.
3. iQC’s hashtable-accelerated fixed-radius KNN search also accelerates “naïve KNN”, because iQC only searches for unique RGBS values. That is, if we hypothetically assume there are infinitely many background pixels in the slide, but all these background pixels have identical red, green, blue, and Sobel magnitude values – then iQC’s hashtable-accelerated fixed-radius KNN will perform exactly one hashtable lookup for that specific red, green, blue, and Sobel magnitude. This single computation is more efficient than a purely naïve KNN, which would compare a pixel to the infinitely many background pixels and never return a result, because there are infinitely many computations to perform.

##### S1.2.12 Write 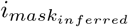

iQC writes 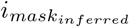 file that includes inferred pixel types.

##### S1.2.13 Biopsy/nonbiopsy prediction

See Surgical procedure prediction (biopsy/nonbiopsy) (Sec S1.4).

##### S1.2.14 Ridge detection

A ridge identifies “window pane breaking” patterns or bubbles. For iQC’s purposes, a ridge is a pixel with an edge type in 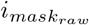. This is a dark/black pixel that has adjacent lighter/background pixels and nearby non-adjacent lighter/background pixels. We consider a Sobel filter to identify edges, and an Otsu on the Sobel magnitudes to classify a pixel type as edge/non-edge. For iQC, an edge pixel is grown outwards to adjacent edge pixels, such that all involved edge pixels are types as ridges. This allows edge pixels that would not themselves be considered ridges to take the ridge type. Canonically in image analysis, a ridge is a contiguous path of edge pixels.

**Fig S1.**
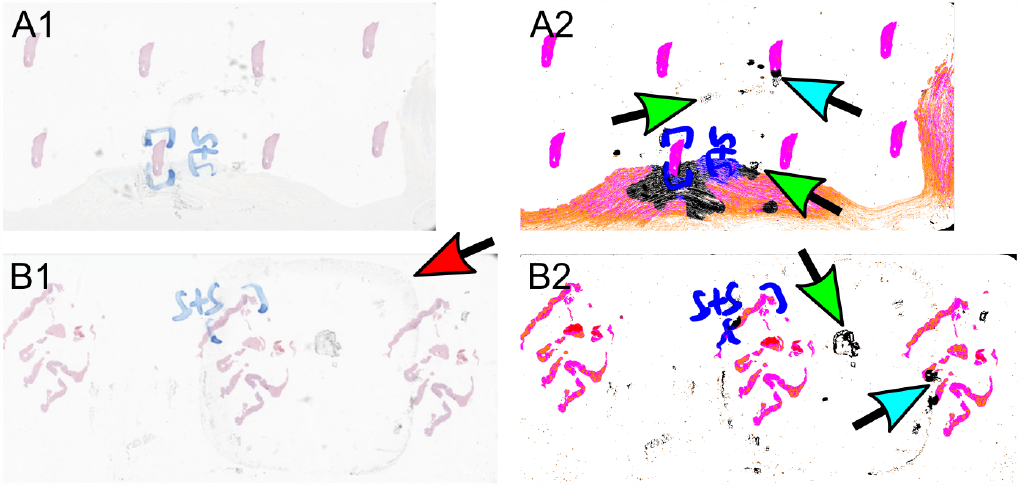
Examples of “window pane breaking” artifacts in VAMC prostate needle biopsy slides. **A1**: Eight levels of a prostate needle biopsy, with extensive artifacts around the bottom and right edges, which may be due to mounting solution leaks and aging. **A2**: A window pane breaking artifact occurs partially over some tissue (*cyan arrow*), and many other window pane breaking artifacts are evident (e.g. at *green arrows*), which may occur as mounting solution breaks down with age. **B1**: Three levels. Additionally, a large square of residue (*red arrow*) may have been deposited on the slide from a physically adjacent sticker. **B2**: A window pane breaking artifact partially overlaps with tissue (*cyan arrow*), and many other window pane breaking artifacts are evident (e.g. at *green arrow*).

###### S1.2.14.1 Acrylamide age statistics, a.k.a. slide degeneration

To estimate how aged/degenerate the slide is, iQC combines ridge detection (edges of bubbles) with stain strength detection (faded stain). More specifically, slide degeneracy metrics take the form of Equation S9:

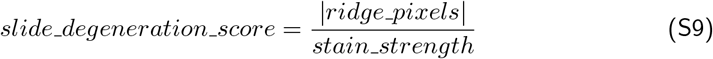

where:

1. |*ridge_pixels*| may be (a) simply the count of ridge pixels, (b) a count of ridge pixels weighted such that ridges pixels farther from the center have a value proportionately less than one in the count, or (c) any other count based on ridge pixels.
2. *stain_strength* may be *stain_strength*_*mean*_ (Eqn S4), *stain_strength*_*median*_ (Eqn S8), or any other metric of stain strength.

In this way, iQC calculates four different *slide_degeneration_score*s, and if at least one of these scores is over 1000, then the slide is “fail_some_tissue” (Sec S1.1). These different *slide_degeneration_score*s are leveraged in the reports iQC builds, so each slide may be ranked according to different criteria, e.g. to identify which slides have the most bubble-related ridges in the center of the slide where the stain may be faded.

##### S1.2.15 Write 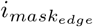

iQC writes 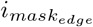 to debug edge and ridge detection. This highlights bubble edges, tissue edges, pen edges, etc (Fig 2C2).

##### S1.2.16 Barcode and PHI/PII detection

Part of the mission at our Center is to make research-ready datasets. It is required that there are no identifiers in research-ready datasets. Identifiers may include PHI/PII as well as medical accession numbers, surgical pathology numbers, etc (Sec 2I1-2).

Therefore, iQC performs barcode detection, which may detect other identifiers printed on a slide as plain text (Sec 2J1-2,K1-2). Barcode detection compares how many dark/black pixels are set against light/background pixels, on both the left and right sides of a slide. If there is a sticker or some printed black text that may disclose identifiers, this text is expected to be on the left side or the right side, but never both sides at the same time. This printed text may be on a black-and-white sticker. Thus, by comparing the dark/light differences on the left to right sides, identifiers such as PHI/PII are most likely to be disclosed when one side has many more dark-against-light pixels than the other side.

##### S1.2.17 Close-out timing statistics

iQC times its performance, for monitoring purposes. These times are reported as iQC completes.

#### S1.3 Lambda operator for contiguity measurements

We define an operator lambda to measure contiguity of pixel types, e.g. how long is an approximately continuous line of tissue type pixels (Alg S1). Some breaks in this line are allowed, but generally the longer the break the lower the score from the lambda operator will be.

##### Algorithm S1

Calculate lamba operator (*λ*) for approximate contiguity of pixel *type* in *mask*, e.g. how many tissue pixel types (*type* = *tissue*) are there approximately in a line of 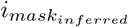. We define *λ*(*mask m, type t*) along the X or Y axis as:

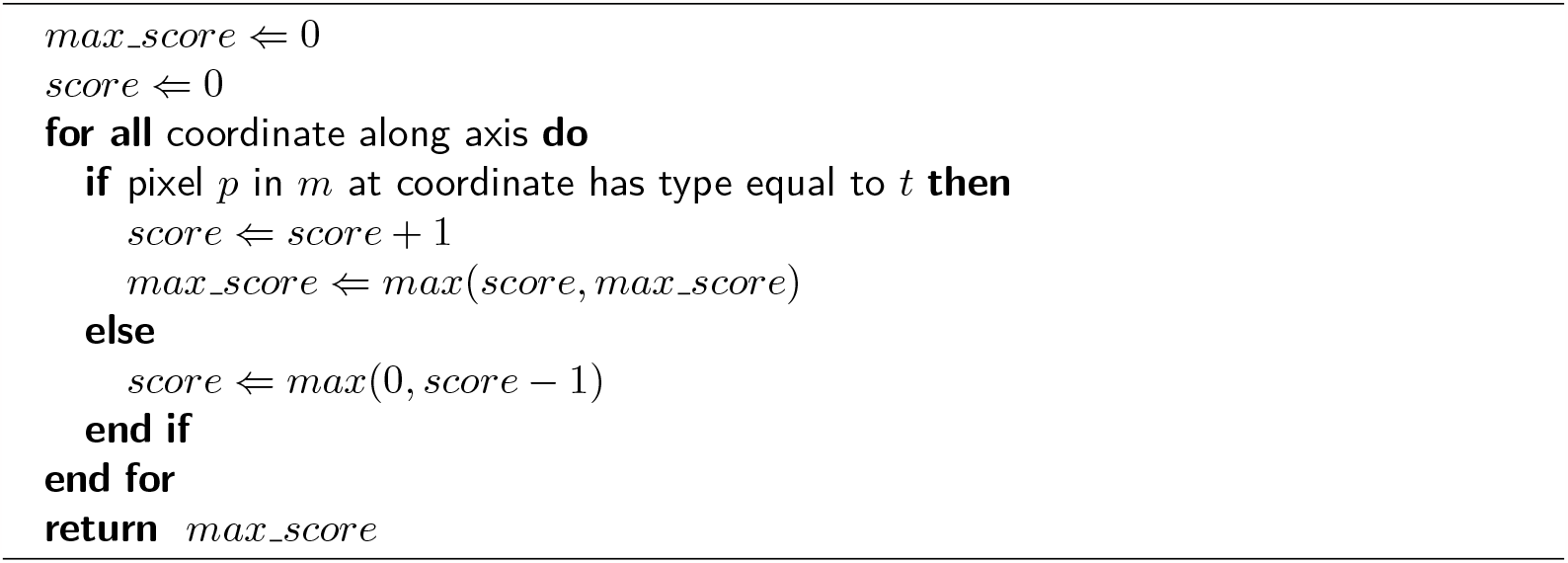

#### S1.4 Surgical procedure prediction (biopsy/nonbiopsy)

In Equation 1, *G*_*narrow*_ converts to a number between 0 and 100 the approximate measurement of the narrowest region of tissue (Eqn S10), with the intuition that biopsies are narrow and will be close to 0:

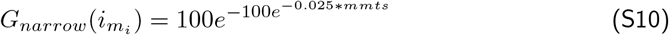

*G*_*area*_ converts to a number between 0 and 100 the approximate measurement of the tissue area by summing up the number of tissue type pixels, with the intuition that biopsies tend to involve little tissue and will be close to 0:

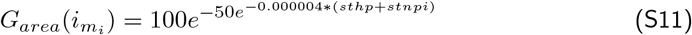

*G*_*long*_ converts to a number between 0 and 100 the approximate length-to-width ratio of the tissue, with the intuition that biopsies tend to have a ratio much greater than 1 (so *G*_*long*_ will be close to 0) while nonbiopsies tend to have a ratio close to 1 (so *G*_*long*_ will be close to 1):

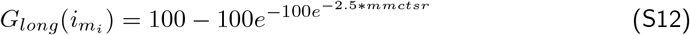

*G*_*adipose*_ is a correction factor for specimens with an abundance of adipose tissue, to prevent some ectomy/TURP samples that are mostly fat from erroneously being predicted as biopsies (Eqn S13). *G*_*adipose*_ converts to a number between 0 and 100000 the approximate measure of how frequently tissue type pixels are adjacent to background type pixels. Background pixels are white/clear/empty in the slide image. Adipose (a.k.a. fat) tissue typically consists of thin strips of stromal tissue to support large globules of fat tissue. The stromal tissue is pink and is counted as tissue pixels, while the fat globules are clear and are counted as background. Therefore in fat, tissue pixels are adjacent to many more background pixels that would occur in a biopsy or in solid blocks of tissue (in most ectomies or TURPs). Fatty ectomies have a high *G*_*adipose*_, which is important because fatty ectomies may have very little solid tissue, or only a thin strip of tissue that would otherwise be classified as a biopsy (e.g. Fig 4B1), if it were not for *G*_*adipose*_ increasing 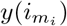 (Eqn 1).

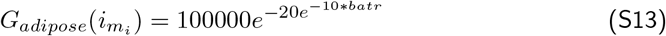

We define *mmts* (Eqn S10) as the “min median tissue score”. Low values of *mmts* mean the narrowest region is very small, so the tissue may be thin, so 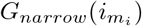 approaches 0, to suggest a biopsy. In contrast, high values of *mmts* mean the narrowest region is much larger, so the tissue is approximately square in shape, 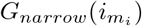 approaches 100, to suggest a nonbiopsy. We calculate *mmts* using lamba operators (Alg S1) across the X and Y axes on 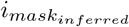 for tissue pixel types.

In Equation S11, we define *sthp* as the “sum [of] tissue or hematoxylin pixels” (which is the total number of pixels in 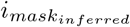 that are tissue or hematoxylin types) and *stnpi* as “suspect-to-nonsuspect pixels inferred” (which is the number of pixels that has the “suspect” type in 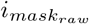 and subsequently were assigned the tissue type in 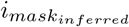).

We define *mmctsr* (Eqn S12) as “maxmin max contiguous tissue score ratio”. Low values of *mmctsr* mean the ratio is close to 1 and the tissue is approximately square so 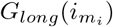 approaches 100, to suggest a nonbiopsy. In contrast, high values of *mmctsr* mean the ratio may be larger than 1 (e.g. a ratio of 20) and the tissue is approximately ribbon-like so 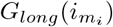 approaches 0, to suggest a biopsy. iQC calculates *mmctsr* as:

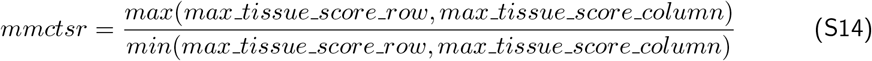

where:

1. *max_tissue_score_row* is calculated using a lamba operator (Alg S1) across the X axis on 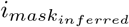 for tissue pixel types.
2. *max_tissue_score_column* is calculated using a lamba operator (Alg S1) across the Y axis on 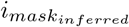 for tissue pixel types.

In Equation S13, we define *batr* as the “background-to-adjacent-tissue ratio”, which is the sum of background-adjacent tissue pixels, divided by the total number of tissue pixels.

#### S1.5 Computational software and hardware

We implemented iQC in `python 3.9.16`, with imports from numpy[32] and `statistics`. Visualization was performed in `R version 4.2.2` and `rstudio 2022.07.2-576`, with plots made via ROCR[33] and ggplot2[34]. Cohen’s *d* calculation[35] and interpretation of D values as negligible(*d <* 0.2)/small(*d <* 0.5)/medium(*d <* 0.8)/large(*d*≥0.8) was performed with R’s effsize package[36]. For the comparison of two independent proportions, Cohen’s *h* calculation[35] was performed with the `ES.h` function of R’s `pwr` package[37]. We calculate effect size in a 2×2 contingency table as *φ* = |*sqrt*(*χ*^2^*/n*)|, where *χ*^2^ is the Chi-squared statistic. For 2*×*2 contingency tables, this *φ* (phi coefficient) is equivalent to Cohen’s *ω* (omega)[35]. The phi coefficient is closely related to the Matthew’s Correlation Coefficient (MCC). We use the `cohenW` function of the R package `rcompanion`[38] to calculate Cohen’s *ω*, for measuring effect size in 2*×*2 contingency tables. We use the same thresholds of “negligible” (effect *<* 0.2), “small” (effect *<* 0.5), “medium” (effect *<* 0.8), and “large” (effect ≥0.8) for Cohen’s *d*, Cohen’s *h*, and *φ* / Cohen’s *ω* effect sizes. For Cohen’s *ω*, Serdar suggests thresholds “negligible” (*ω <* 0.1), “small” (*ω <* 0.3), “medium” (*ω <* 0.5), and “large” (*ω*≥0.5), but these thresholds do not change our Association findings shown in the bottom row of Table 5. Text processing was performed in `perl 5.32.1`.

For computation, we leveraged the Genomic Information System for Integrative Science (GenISIS) datacenter at the Center for Data and Computational Sciences. iQC can run on a single-CPU system with an amount of CPU RAM approximately triple the file size, e.g. for a 1GB whole slide image, we suggest 3GB CPU RAM. We recommend running iQC on at least 5 CPUs in parallel, ideally 20 CPUs, and 80+ CPUs for best performance. The amount of required CPU RAM scales with the number of parallel CPUs.

For comparisons to iQC, we considered HistoQC[6] at commit version a99916c2ceaa61c7ae5d75600f10d3fb553041fc from March 22, 2023. We used HistoQC’s default configuration file, i.e. config.ini. We disabled the machine-learning-driven coverslip and pen detection modules in our custom configuration file for HistoQC per tickets 250, 254, and 269 from April to November 2023.

#### S1.6 Glass microscopy slide preparation

Institutions vary in slide preparation protocols, e.g. mounting media choice and automated coverslip instrument. Institutions had the same slide staining instrument, i.e. Tissue-Tek Automated Slide Stainer, model Prisma Plus (A-10), manufactured by Sakura USA.

##### S1.6.1 Mounting Media and Coverslip Instrument differ among Institutions

Institution *α*’s pathology lab prepared glass microscopy slides with Acrymount Plus (StatLab Medical Products, McKinney, TX), a Toluene and Acrylate polymer based mounting media (a.k.a. “mounting solution”). This reagent is formulated to reduce oxidation, discoloration (i.e. yellowing) and fading in H&E stains over time. This may be superior in quality and integrity to the more traditional Xylene-based mounting media. Additionally, Institution *α* used a Tissue-Tek Automated Glass Coverslipper, model Glas g2, manufactured by Sakura USA.

Instead of mounting media, Institution *β*’s pathology lab prepared glass microscopy slides with a resin-coated plastic film, a.k.a. “coverslip tape” or “mounting tape.” Specifically, this is Tissue-Tek Coverslipping Film, Cat# 4770, manufactured by Sakura USA. Additionally, Institution *β* used a Tissue-Tek Film Coverslipper, model 4740, manufactured by Sakura USA.

We suspect the proper mounting media plays a critical role in embedding the specimen as well as providing an optically translucent barrier that preserves the pathological tissue section quality and integrity for long-term storage. Furthermore, a high-quality mounting media will have a refractive index that is virtually identical to the glass slide. This may be optimal for high quality images and high magnification[39].

Leakage around slides (Fig S1A1) is usually attributed to usage of excessive amount of mounting media or low-quality ingredients within the media that may degrade over time and form byproducts, e.g. acrylamide bubbles or “window pane breaking” artifacts.

#### S1.7 Whole slide images

VAMC glass microscopy slides were scanned on a Leica Aperio GT450 scanner, which produces SVS files that we read via `openslide 3.4.1`[40, 41]. AGGC slides were scanned on an Akoya Biosciences scanner, which produces TIFF files that we read via `ImageMagick 6`.

#### S1.8 Machine learning

iQC uses machine learning to infer select “suspect” pixel types to other pixel types. Specifically, for a given image, iQC uses the K-nearest neighbors algorithm[42, 43], with k=1 and an L1 norm. A pixel is represented as a four-dimensional value: red channel (0 to 255 integer), green channel, blue channel, and Sobel channel. The Sobel channel is the magnitude of a 3 *×* 3 Sobel operator[44, 45].

#### S1.9 Workflow icons attribution

For Figure 1, we leverage a number of icons from The Noun Project^1^ (Table S1).

**Table S1.**
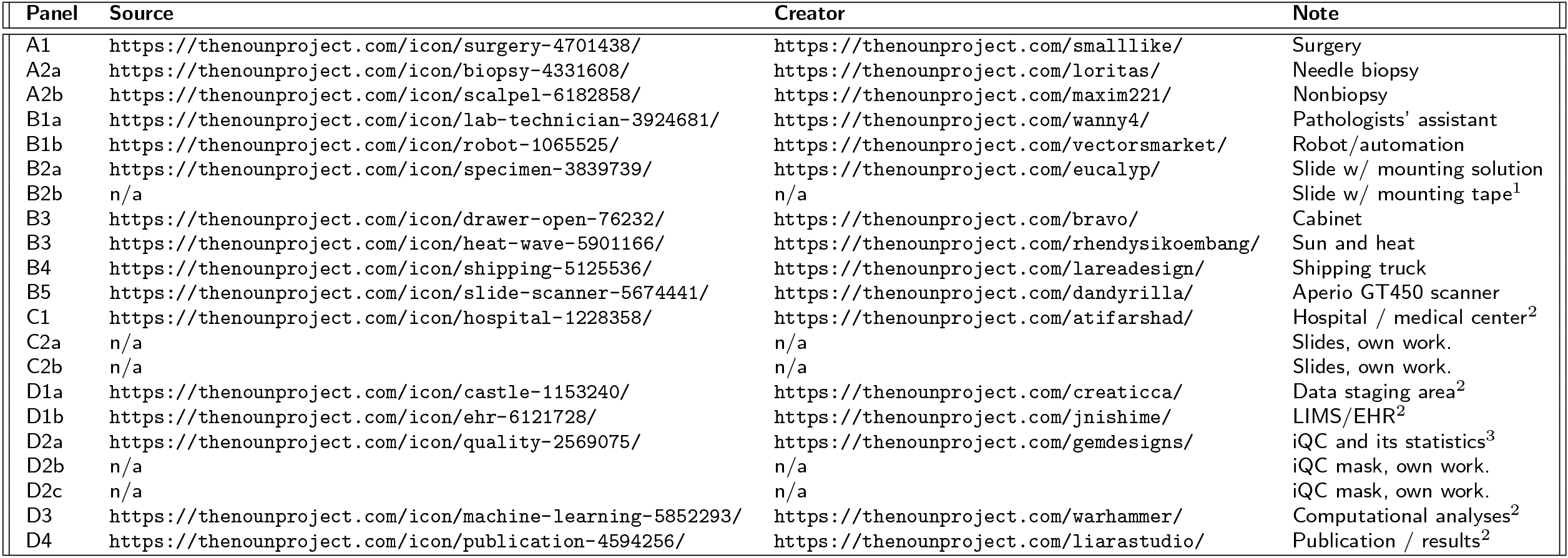
Attribution of icons in Figure 1 panels. Icons are distributed under a Creative Commons Attribution 3.0 license https://creativecommons.org/licenses/by/3.0/. Legend: ^1^ is own/AJS work, derived from B2a. ^2^ color is own/AJS work. ^3^ text, color, and background are own/AJS work.

## Footnotes

1 The Noun Project may be reached at https://thenounproject.com

